# Prediction of Central Post-Stroke Pain by Quantitative Sensory Testing

**DOI:** 10.1101/2024.04.24.24305954

**Authors:** Susanna Asseyer, Eleni Panagoulas, Jana Maidhof, Kersten Villringer, Esra Al, Xiuhui Chen, Thomas Krause, Samyogita Hardikar, Arno Villringer, Gerhard Jan Jungehülsing

## Abstract

**Objective:** Among patients with acute stroke, we aimed to identify those who will later develop central post-stroke pain (CPSP) versus those who will not (non-pain sensory stroke: NPSS) by assessing potential differences in somatosensory profile patterns and evaluating their potential as predictors of CPSP.

**Methods:** We performed a prospective longitudinal quantitative sensory testing (QST) study in 75 stroke patients with somatosensory symptoms, recruited in the acute phase and followed up for 12 months. Based on previous QST studies in chronic stroke, we hypothesised that QST values of cold detection threshold (CDT) and dynamic mechanical allodynia (DMA) would differ between CPSP and NPSS patients before the onset of pain. Mann-Whitney U-tests and mixed ANOVAs with Bonferroni corrections were performed to compare z-normalised QST scores between both groups.

**Results:** In total 26 patients (34.7%) developed CPSP. In the acute phase, CPSP patients showed significant contralesional cold hypoesthesia compared to NPSS patients (*P* = 0.04), but no significant DMA differences. Additional exploratory analysis showed NPSS patients exhibit cold hyperalgesia on the contralesional side compared to the ipsilesional side, not seen in CPSP patients (*P* = 0.011). A gradient-boosting approach to predicting CPSP from QST patterns prior to pain onset, had an overall accuracy of 84.6, with a recall and precision of 0.75. Notably, both in the acute and the chronic phase, about 80% of CPSP and NPSS patients showed bilateral QST abnormalities.

**Interpretation:** Cold perception differences between CPSP and NPSS patients appear early post stroke before pain. Prediction of CPSP through QST patterns seems feasible.

## Introduction

Central post-stroke pain (CPSP) is a severe form of neuropathic pain that is often refractory to treatment^1^ affecting about 8% to 10.5 % of patients after stroke.^2,3^ It occurs within weeks to months in patients with lesions in the central somatosensory system, including the thalamus^4–7^, brainstem, pons, and somatosensory cortices.^8^ In patients with thalamic stroke prevalence has been reported to be 18% and in those with a lateral medullary infarction 25%,^9,10^ though reported estimates of prevalence vary widely. Pain localization is typically associated with sensory abnormalities in body parts corresponding to the affected brain area.^1^ Besides spontaneous and/or evoked pain, impaired temperature sensation and nociception have been reported.^11^ This has led to the assumption that lesions of the lateral spinothalamic tract (STT) and/or its central projections to the cortex are a prerequisite for the development of CPSP.^12–14^ Consistent with this assumption, studies have shown differences in the location of thalamic lesions in patients with and without pain, with lesions in CPSP patients often involving the ventral posterior nucleus (VPN) and the anterior pulvinar nucleus, where the STT is thought to terminate.^5–7,15^

Few studies have examined patients in the acute phase after stroke before the onset of pain.^2,9,11^ Klit *et al.*^11^ reported in acute stroke patients that a combination of reduced sensation to pinprick or cold, and evoked pain or dysesthesia, increases the risk of developing CPSP, when comparing the affected to the unaffected body parts.^11^ What has been missing is a standardised and quantifiable assessment of clinical and specific sensory symptoms in the pre-pain phase, which would allow an easier transfer of results to clinical practice.

To address this need, we conducted a prospective longitudinal study in patients with acute somatosensory stroke as part of a larger prospective clinical trial^16^ in which, in addition to a detailed clinical assessment and MR imaging, standardised quantitative sensory testing (QST) was performed.^17^ QST was conducted in the acute/subacute phase “before pain” and follow-up exams were repeated at different time points up to 12 months.

Based on two previous QST studies in *chronic* CPSP patients,^18,19^ we hypothesized that the QST parameters, cold detection threshold (CDT) and dynamic mechanical allodynia (DMA), differ already in the *acute* stage of stroke between patients who will later develop CPSP and those who will not (NPSS: Non Pain Sensory Stroke). In addition, we performed an exploratory analysis of all QST parameters bilaterally in the acute and chronic phases after stroke.

## Materials and methods

### Subjects

Patients with an acute ischemic or haemorrhagic stroke affecting the somatosensory system were enrolled at the Charité-Universitätsmedizin Berlin, Germany between 2010 and 2016. The study was approved by the local ethics committee (EA4/003/10) and informed consent was obtained from all participants prior to inclusion.

Inclusion criteria were age between 18 and 85 years and a transient or persistent somatosensory deficit with MRI-proven acute unilateral stroke within central parts of the somatosensory system, i.e., medulla oblongata, pons, thalamus, internal capsule, or somatosensory cortices Exclusion criteria can be seen in the supplementary methods. Patients were allocated to the two groups CPSP or NPSS according to their outcome regarding pain.

Group classification was based on medical history, neurological examination, and responses to pain questionnaires.

CPSP was defined as pain or unpleasant sensation that occurred as a direct result of a stroke to the central somatosensory system in the body area corresponding to the somatosensory deficit (e.g., hypoesthesia, paraesthesia).^1,20^ CPSP most commonly occurs within the first few months after stroke^2,11,21^ therefore, in-person follow up for the diagnosis of CPSP was chosen to be at least six months after stroke.

### Clinical data and assessment

Each patient underwent a semi-structured interview for medical history and a neurological examination (see supplementary methods) including stroke severity assessment (National Institutes of Health Stroke Scale (NIHSS)^22^, modified Rankin Scale (mRS)^23^, and Barthel Index (BI)^24^) as well as QST, and MRI. Patient-reported outcomes were assessed by validated questionnaires (12-item Short Form Health Survey (SF-12)^25^, Pittsburgh Sleep Quality Index (PSQI)^26^, Geriatric Depression Scale 30 (GDS-30)^27^). For details of the study protocol see supplementary Table S1A and B. Only in patients who reported pain, the painDETECT Questionnaire (PD-Q), the German pain questionnaire (DSF) and pain perception scale (SES) were administered.^28–30^

### Quantitative sensory testing

Our study aimed to capture QST data in the acute/subacute stage before pain development to identify potential differences between CPSP and NPSS patients. QST is a standardized, non-invasive method sanctioned by the DFNS,^31^ which evaluates somatosensory function using normative values from a healthy population, adjusted for age, sex, and test location.^17^ Examinations targeted the somatosensory system’s most affected area (face, hand, or foot), covering both ipsilesional and contralesional sides, in sequence. QST parameters are denoted as “_c_” for contralesional (e.g., _c_CDT) and “_i_” for ipsilesional (e.g., _i_CDT), with side-to-side differences labelled “_sd_” (e.g., _sd_CDT), calculated by subtracting ipsilesional from contralesional values. All assessments were conducted by two DFNS-trained examiners.

QST determines the following parameters: Cold (CDT) and warm detection threshold (WDT), thermal sensory limen (TSL), paradoxical heat sensations (PHS), cold (CPT) and heat pain threshold (HPT), mechanical detection threshold (MDT), mechanical pain threshold (MPT), mechanical pain sensitivity (MPS), DMA, wind-up ratio (WUR), vibration detection threshold (VDT), and pressure pain threshold (PPT).^17^

### Statistical analysis

Statistical analysis was conducted in R 4.2.2 (2022.10.31)^32^ with RStudio, data normality was assessed through skewness, kurtosis, histograms, and the Shapiro-Wilk test. Normally distributed metrics are reported as mean ± SD, and non-normally distributed as median and range. Group comparisons (CPSP vs. NPSS) utilized Pearson’s chi-square for categorical variables and the Mann-Whitney U test for non-normally distributed ordinal/quantitative data, with Cliff’s delta indicating effect size. Comparison between patients and the reference values from the healthy reference collective was conducted as previously suggested.^33^ See supplementary methods for more details. Significance was set at p ≤ .05.

Longitudinal data analysis, addressing non-normality and outliers, applied robust statistical methods from the WRS2 (1.1-4) package in R,^34^ including median-based imputation for missing values. A mixed-design was evaluated using robust ANOVA (bwtrim), focusing on within- and between-subject effects, with post hoc analysis (sppbi) exploring “Group” and “Time” interactions. Robustness was further ensured by employing the “mom” M-estimator for individual contrasts and bootstrap resampling (nboot=10000) to validate findings. Logistic regression was performed to assess possible indicators for the development of CPSP. The *finalfit* (1.0.7) package was used in R to produce the final regression tables and odds ratio figures.^35^ Binary logistic regression examined various variables (age, sex, neurological scores—BI, mRS, NIHSS, health quality, QST) independently. In a second step, significant variables from the binary regression were selected and multiple logistic regression was performed. Model efficacy was evaluated using Nagelkerke’s R², with model comparison based on Akaike Information Criterion (AIC) and C-statistic. To avoid overlapping parameters in the multiple regression analysis, NIHSS was selected as a parameter representative for neurological impairment.

### Prediction of CPSP

Python (3.11.5) and scikit-learn^36^ were used to train a GradientBoostingClassifier algorithm to classify patients before occurrence of pain into CPSP and NPSS groups using the QST parameters. Leave-one-out-cross-validation (LOOCV) was used to validate the model. All QST variables from the contralesional side and side-to-side difference were initially included in the classifier. In the final classifier QST variables were chosen based on feature importance and those that were significantly different between CPSP and NPSS patients. The hyperparameters of the algorithm were optimised using GridSearchCV. The GridSearchCV object was configured with the GradientBoostingClassifier as the base estimator, and the evaluation metric was set to the weighted F1 score. LOOCV was used to ensure robust model assessment. The optimal configuration, comprising 180 trees, a learning rate of 0.1, ‘log_loss’ as the loss function, and a maximum tree depth of 3, was then used for the final classifier. The accuracy, recall, precision, ROC-AUC, F1 scores, and confusion matrix of this model are reported. Furthermore, permutation importance was used to determine which QST variables contributed the most in the classification of pain patients.

### Data availability

Data are available upon reasonable request. The analysis code will be made available on github, epanagoulas/SOSENS

## Results

### Cohort description

Of 115 patients screened after acute unilateral somatosensory stroke, 75 were included; 26 developed CPSP and 49 did not (NPSS) (Figure 1). Six CPSP patients (incomplete data or pain before first QST) and four NPSS patients (lacking early QST data) were excluded from the pre-pain and prospective QST. The final QST analysis involved 20 CPSP and 45 NPSS patients. For the hypothesis driven analysis focusing on pre-pain QST, 18 CPSP and 38 NPSS patients were evaluated, excluding nine subjects previously published to prevent data overlap.^18^ An overview of clinical characteristics and results of questionnaires is given in Table 1.

**Figure 1:**
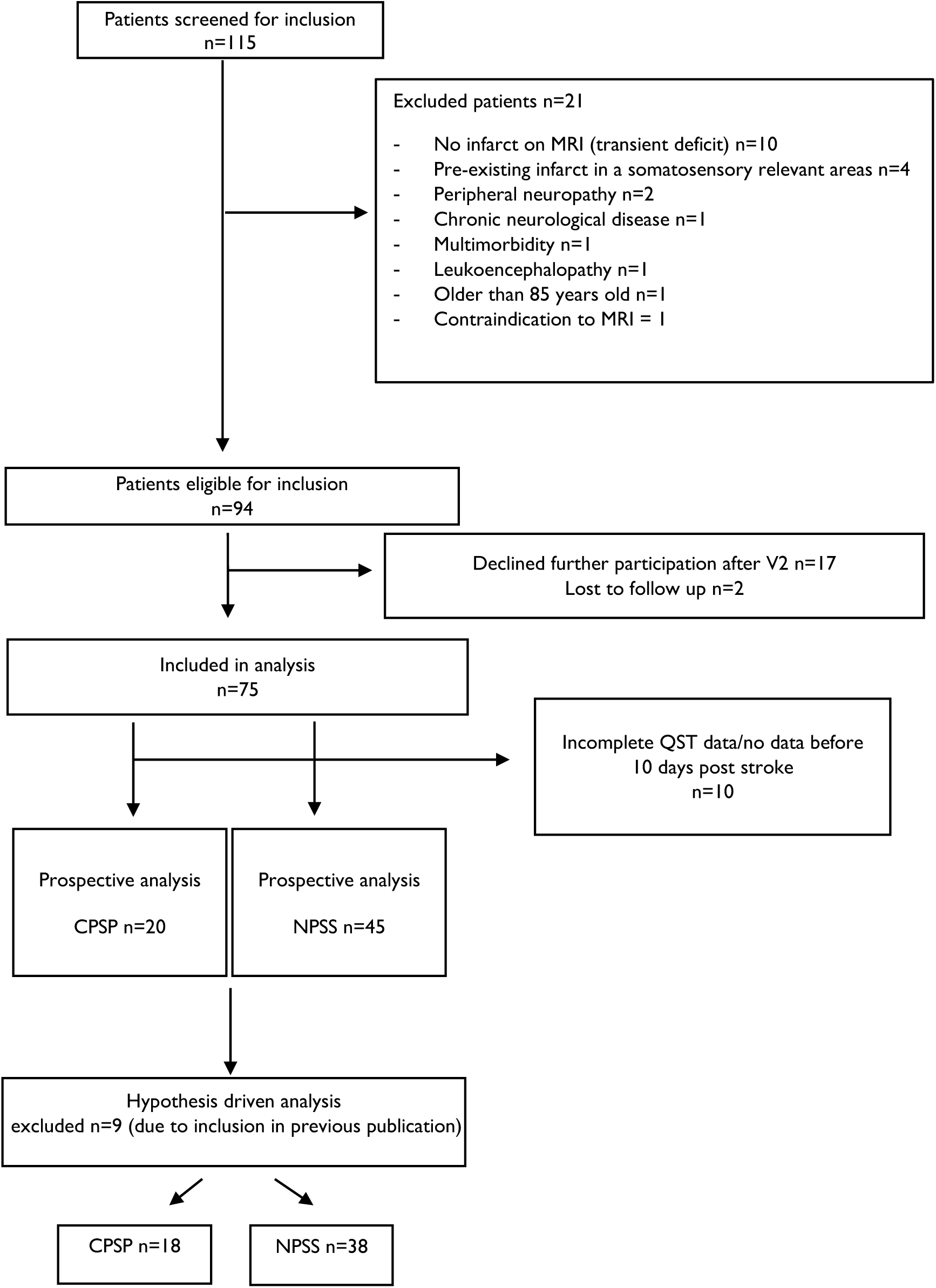
Patient recruitment

**Table 1:** Demographics and Clinical Data.

| Diagnosis |  | CPSP | NPSS | Total | P |
| --- | --- | --- | --- | --- | --- |
| Total | N (%) | 26 (34.7) | 49 (65.3) | 75 |  |
| Age (years) | Median (IQR) | 63.0 (55.5 to 69.5) | 65.0 (56.0 to 70.0) | 65.0 (55.5 to 70.0) | 0.660 |
| Sex | Female / N (%) | 15 (57.7) | 15 (30.6) | 30 (40.0) | **0.028** |
|  | Male / N (%) | 11 (42.3) | 34 (69.4) | 45 (60.0) |  |
| Thrombolysis | No / N (%) | 18 (69.2) | 40 (81.6) | 58 (77.3) | 0.255 |
|  | Yes / N (%) | 8 (30.8) | 9 (18.4) | 17 (22.7) |  |
| Aetiology | Haemorrhagic / N (%) | 1 (3.8) | 2 (4.1) | 3 (4.0) | 1.000 |
|  | Ischemic / N (%) | 25 (96.2) | 47 (95.9) | 72 (96.0) |  |
| Lesion side | Left / N (%) | 9 (34.6) | 24 (49.0) | 33 (44.0) | 0.329 |
|  | Right / N (%) | 17 (65.4) | 25 (51.0) | 42 (56.0) |  |
| Lesion location | Brainstem / N (%) | 6 (23.1) | 5 (10.2) | 11 (14.7) | 0.191 |
|  | Cortex / N (%) | 7 (26.9) | 8 (16.3) | 15 (20.0) |  |
|  | Thalamus / N (%) | 13 (50.0) | 35 (71.4) | 48 (64.0) |  |
|  | Pathways / N (%) |  | 1 (2.0) | 1 (1.3) |  |
| Hypertension | no / N (%) | 8 (30.8) | 8 (16.3) | 16 (21.3) | 0.235 |
|  | yes / N (%) | 18 (69.2) | 41 (83.7) | 59 (78.7) |  |
| Diabetes | no / N (%) | 23 (88.5) | 42 (85.7) | 65 (86.7) | 1.000 |
|  | yes / N (%) | 3 (11.5) | 7 (14.3) | 10 (13.3) |  |
| Smoking | no / N (%) | 19 (73.1) | 35 (71.4) | 54 (72.0) | 1.000 |
|  | yes / N (%) | 7 (26.9) | 14 (28.6) | 21 (28.0) |  |
| Cholesterol | no / N (%) | 2 (7.7) | 16 (32.7) | 18 (24.0) | **0.022** |
|  | yes / N (%) | 24 (92.3) | 33 (67.3) | 57 (76.0) |  |
| Atrial fibrillation | no / N (%) | 22 (84.6) | 43 (87.8) | 65 (86.7) | 0.731 |
|  | yes / N (%) | 4 (15.4) | 6 (12.2) | 10 (13.3) |  |
| Obesity (BMI >30) | no / N (%) | 22 (84.6) | 46 (93.9) | 68 (90.7) | 0.227 |
|  | yes / N (%) | 4 (15.4) | 3 (6.1) | 7 (9.3) |  |
| Family history CVD | no / N (%) | 25 (96.2) | 42 (85.7) | 67 (89.3) | 0.249 |
|  | yes / N (%) | 1 (3.8) | 7 (14.3) | 8 (10.7) |  |
| Assessment scales |  | score |  |  |  |
| First NIHSS | Median (IQR) | 3.0 (2.0 to 5.0) | 2.0 (1.0 to 2.0) | 2.0 (1.0 to 3.0) | **0.001** |
| First mRS | Median (IQR) | 2.0 (1.0 to 3.8) | 1.0 (1.0 to 2.0) | 1.0 (1.0 to 2.0) | **0.001** |
| First Barthel index | Median (IQR) | 90.0 (61.2 to 100.0) | 100.0 (100.0 to 100.0) | 100.0 (90.0 to 100.0) | **0.002** |
| Chronic NIHSS | Median (IQR) | 1.0 (1.0 to 2.0) | 1.0 (0.0 to 1.0) | 1.0 (0.0 to 1.0) | **0.001** |
| Chronic mRS | Median (IQR) | 1.0 (1.0 to 2.0) | 1.0 (1.0 to 1.0) | 1.0 (1.0 to 1.0) | **<0.001** |
| Chronic Barthel index | Median (IQR) | 100.0 (100.0 to 100.0) | 100.0 (100.0 to 100.0) | 100.0 (100.0 to 100.0) | 0.239 |
| Acute PSQI | Median (IQR) | 6.0 (3.0 to 7.0) | 4.0 (2.0 to 6.0) | 5.0 (3.0 to 7.0) | 0.078 |
| Chronic PSQI | Median (IQR) | 6.0 (3.2 to 8.8) | 4.0 (3.0 to 5.0) | 4.0 (3.0 to 6.5) | **0.019** |
| Acute GDS | Median (IQR) | 5.0 (2.0 to 8.0) | 4.0 (1.0 to 7.0) | 4.0 (1.0 to 8.0) | 0.557 |
| Chronic GDS | Median (IQR) | 4.5 (2.0 to 7.8) | 4.0 (2.0 to 7.0) | 4.0 (2.0 to 7.0) | 0.447 |
| Acute SF-12-MCS | Median (IQR) | 55.8 (48.4 to 60.8) | 55.9 (50.7 to 58.4) | 55.9 (50.3 to 59.0) | 0.504 |
| Acute SF-12-PCS | Median (IQR) | 45.0 (42.5 to 52.9) | 51.7 (47.4 to 54.2) | 50.3 (44.6 to 54.0) | **0.025** |
| Chronic SF-12-MCS | Median (IQR) | 55.9 (53.5 to 58.8) | 55.9 (50.2 to 57.8) | 55.9 (51.4 to 58.2) | 0.581 |
| Chronic SF-12-PCS | Median (IQR) | 50.2 (40.7 to 53.8) | 53.6 (47.2 to 55.5) | 53.1 (45.6 to 54.6) | **0.021** |
| Pain occurrence within |  |  |  |  |  |
| <= 1 week | N (%) | 5 (19.2) | 0 |  |  |
| >1 week to <= 1 month | N (%) | 8 (30.8) | 0 |  |  |
| > 1 month <= 3 months | N (%) | 7 (26.9) | 0 |  |  |
| > 3 months <= 8 months | N (%) | 6 (23.1) | 0 |  |  |

CPSP, Central post-stroke pain; NPSS, Non-pain sensory stroke; BMI, body mass index; CPSP, Central post-stroke pain; CVD, cardiovascular disease, NIHSS, national institutes of health stroke scale; mRS, modified Rankin scale, PSQI, Pittsburgh sleep quality index, GDS, geriatric depression scale, SF-12-MCS, short form 12 mental component score, SF-12-PCS, short form 12 physical component score.

Sensory deficits were localised unilaterally in the body and/or face on the contralesional side to the stroke. Results of the non-quantitative clinical examination of sensory symptoms are reported in supplementary Table S2.

### Pain features

Patients developed CPSP within 8 months (mean=60.6, SD=64.2 days) following stroke, detailed analysis of pain onset is reported in Table 1. Pain localisation is displayed in Figure 2. Average pain intensity was 4.1 ±1.9 (SD) and maximum pain intensity was 6.3 ± 2.1 (mean ± SD) from a scale of 0 to 10 (worst pain imaginable). The mean PD-Q score was 12.4 ± 6.7 (SD). Common pain descriptors included burning (*n=*12), pressing (*n=*9), stinging (*n=*8), throbbing (*n=*8) or knocking (*n=*7), severe (*n*=10) and annoying (*n=*9).

**Figure 2.**
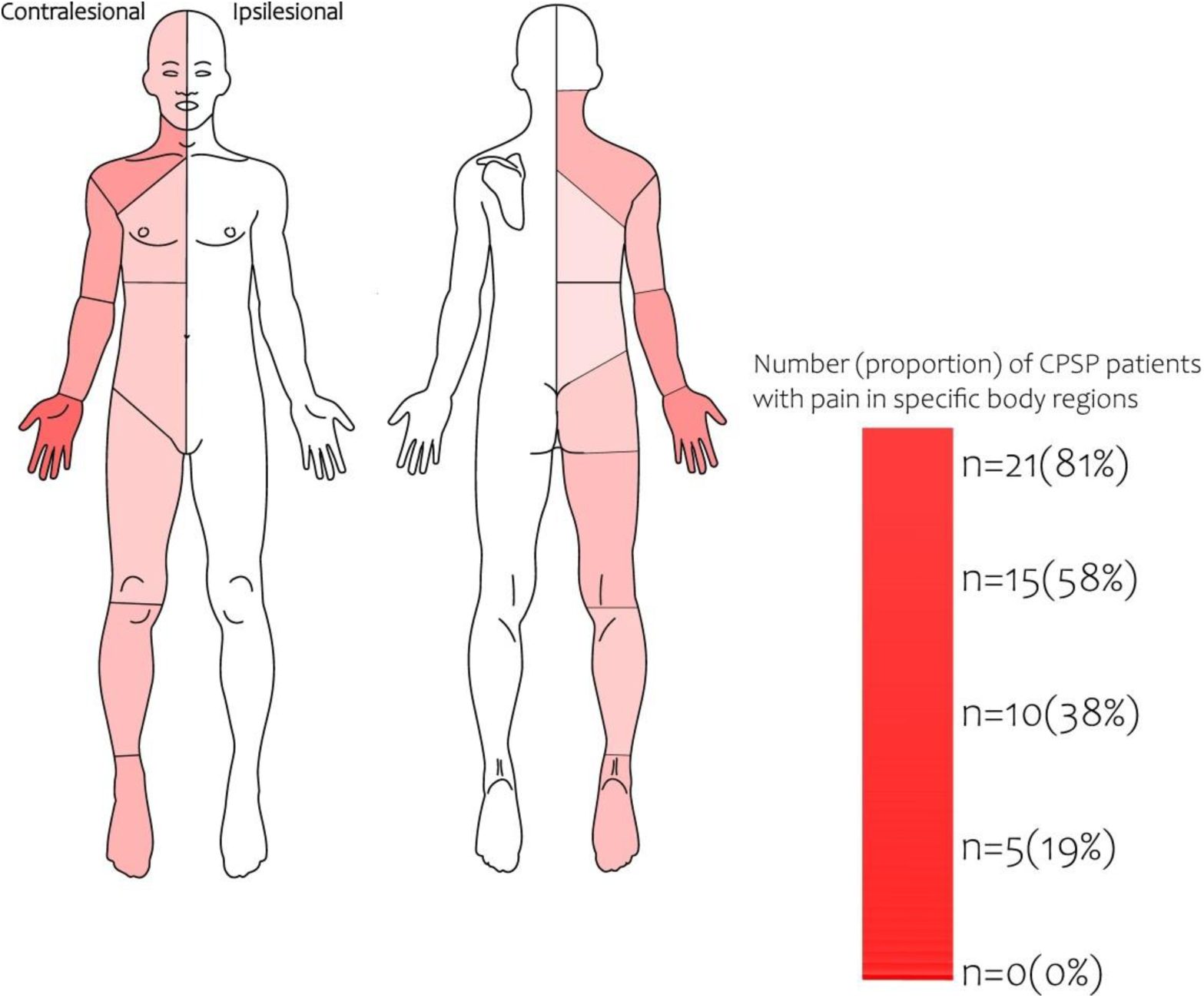
Pain localisation.

Flow chart of prospective patient recruitment. NPSS=Non pain sensory stroke, CPSP=Central post-stroke pain.

Pain localisation of patients with central post stroke pain (CPSP) (n=26). All unilateral right-sided infarcts with clinical symptoms have been flipped to the left side so that all symptoms are depicted on the same body side. Pain contralesional to the stroke lesion manifested in the face (n=4), perioral region (n=8), shoulder and upper arm (n=10), forearm (n=12), hand (n=21),chest and abdomen (n=3), buttock (n=5), thigh (n=4), lower leg (n=7) and foot (n=8).

### Quantitative sensory testing

The hypothesis driven group comparisons and the exploratory group comparisons are given below. The analyses of individual QST values compared to the DFNS reference collective are given in the supplementary Tables S4, S5, and Fig. S1 and described in the supplementary results.

QST in the “acute phase” of stroke before eventual pain

QST was performed within 1-10 days after stroke (mean*=*3.9, SD=1.97 days

#### CDT and DMA (hypothesis driven comparison)

Based on the aforementioned hypotheses we expected CPSP patients to show differences in CDT and DMA compared to NPSS patients before pain occurred. Indeed, CPSP patients showed contralesional hypoesthesia to cold compared to NPSS patients (_c_CDT *U*=475, *P*=0.04 Bonferroni corrected) with a medium effect size Cliff’s delta=-0.39 (Table 2). This confirms that NPSS patients had a consistently lower cold detection threshold (i.e., higher z scores) than CPSP patients. DMA did not differ between the two groups (*U*=368, *P=*0.54).

**Table 2.** Hypothesis driven analysis of QST parameters.

| Diagnosis |  | CPSP | NPSS | P | P adj | d |
| --- | --- | --- | --- | --- | --- | --- |
| Acute setting |  |  |  |  |  |  |
| Total N (%) |  | 18 (32.1) | 38 (67.9) |  |  |  |
| cCDT | Median (IQR) | -1.5 (-3.5 to -0.5) | -0.2 (-1.3 to 0.2) | 0.020 | **0.04** | **-0.39** |
| cDMA | Median (IQR) | 0.0 (0.0 to 0.0) | 0.0 (0.0 to 0.0) | 0.538 | 1.00 |  |

CPSP, Central post-stroke pain; NPSS, Non-pain sensory stroke; CDT, cold detection threshold; DMA, dynamic mechanical allodynia; QST on the contralesional side is indicated with “c” (e.g., cCDT).

##### Exploratory comparisons of all QST parameters in the pre-pain phase

- **Contralesionally**, CPSP patients exhibited pronounced thermal hypoesthesia (_c_CDT *U*=631, *P=*0.010; _c_TSL *U*=615, *P*=0.019) and hypoalgesia (_c_CPT *U*=643, *P*=0.006; _c_HPT *U*=565, *P=*0.043) compared to NPSS patients, with significant differences in cold, warmth, and pressure (_c_PPT *U*=608, *P*=0.005). NPSS patients showed a mechanical hyperalgesia compared to CPSP patients (_c_MPS *U*=589, *P*=0.048).
- **Ipsilesionally**, pallesthesia differed between CPSP and NPSS patients (iVDT U=594, P=0.039), with more pronounced pallhypoesthesia in patients with CPSP.
- A marked **side-to-side difference** in cold pain threshold (_sd_CPT *U*=682, *P* < 0.001) with a large effect size Cliff’s *d*=-0.55 indicated NPSS patients experienced heightened sensitivity to cold pain on the contralesional side, contrasting with CPSP patients’ preserved thresholds. CPSP patients also showed a relative heat pain hypoalgesia on the contralesional side compared to non-pain patients *U*=586, *P*=0.012 (Cliff’s *d*=-0.40 medium effect size) and a relative loss of warm cold differentiation (TSL) on the contralesional side *U*=590, *P=*0.03 (Cliff’s *d*=-0.34, medium). Only _sd_CPT survived Bonferroni correction (_sd_CPT *P*=0.0011).

Results are detailed in Table 3-5a and supplementary Fig. S2A-C.

**Table 3.** QST parameters in acute (pre-pain) phase and chronic phase: contralesional side.

|  |  | Between group comparison |  |  |  | Between group comparison |  |  |  |
| --- | --- | --- | --- | --- | --- | --- | --- | --- | --- |
| Diagnosis |  | a. Acute |  | P | d | b. Chronic |  | P | d |
|  |  | CPSP | NPSS |  |  | CPSP | NPSS |  |  |
| Total N (%) |  | 20 (30.8) | 45 (69.2) |  |  | 20 (30.8) | 45 (69.2) |  |  |
| cCDT | Median (IQR) | -1.3 (-3.0 to -0.4) | -0.2 (-1.1 to 0.2) | **0.010\*** | -0.40 | -0.8 (-2.5 to -0.3) | -0.1 (-0.8 to 0.4) | **0.007\*** | -0.40 |
| cWDT | Median (IQR) | -1.8 (-2.9 to -0.6) | -0.7 (-1.9 to 0.0) | 0.059 |  | -2.2 (-2.9 to -0.7) | -0.6 (-1.5 to 0.4) | **0.014\*** | -0.30 |
| cTSL | Median (IQR) | -1.4 (-2.5 to -0.7) | -0.7 (-1.4 to 0.2) | **0.019\*** | -0.37 | -1.7 (-2.4 to 0.1) | -0.6 (-1.1 to 0.1) | 0.064 |  |
| cCPT | Median (IQR) | -0.5 (-0.9 to 0.7) | 1.2 (-0.4 to 2.2) | **0.006\*** | -0.43 | -0.3 (-0.9 to 1.4) | 1.0 (-0.2 to 2.0) | **0.030\*** | -0.43 |
| cHPT | Median (IQR) | -1.2 (-1.5 to -0.5) | -0.1 (-1.1 to 0.9) | **0.043\*** | -0.32 | -0.9 (-1.3 to 0.6) | 0.0 (-0.9 to 1.5) | **0.046\*** | -0.32 |
| cPPT | Median (IQR) | -1.2 (-2.4 to -0.6) | -0.3 (-1.2 to 0.5) | **0.005\*** | -0.45 | -1.2 (-1.9 to 0.4) | -0.5 (-1.2 to 0.5) | 0.053 |  |
| cMPT | Median (IQR) | 0.6 (-0.9 to 2.4) | 1.2 (0.0 to 2.3) | 0.309 |  | 1.5 (-0.7 to 3.4) | 1.7 (0.4 to 2.8) | 0.446 |  |
| cMPS | Median (IQR) | 0.2 (-1.1 to 1.5) | 1.3 (0.1 to 2.3) | **0.048\*** | -0.31 | 1.4 (-0.3 to 2.0) | 1.2 (0.5 to 2.5) | 0.233 |  |
| cWUR | Median (IQR) | 0.4 (-0.4 to 0.7) | -0.1 (-0.7 to 0.5) | 0.564 |  | 0.0 (-0.6 to 0.8) | -0.3 (-0.7 to 0.6) | 0.649 |  |
| cMDT | Median (IQR) | -1.6 (-2.9 to 0.3) | -0.7 (-2.7 to 0.0) | 0.546 |  | -1.0 (-1.9 to -0.4) | -0.3 (-1.5 to 0.6) | **0.047\*** | -0.09 |
| cVDT | Median (IQR) | -0.5 (-1.2 to 0.8) | 0.8 (-1.0 to 0.9) | 0.089 |  | 0.1 (-1.1 to 0.5) | 0.8 (0.1 to 0.9) | **0.010\*** | -0.26 |
| cPHS | 0 | 16 (80.0) | 34 (75.6) | 0.217 |  | 16(84.2) | 42(97.7) | 0.082 |  |
|  | 1 | 1 (5.0) | 7 (15.6) |  |  | 1(5.3) | 0(0.0) |  |  |
|  | 2 | 1 (5.0) | 0 (0.0) |  |  | 0(0.0) | 0(0.0) |  |  |
|  | 3 | 0 (0.0) | 3 (6.7) |  |  | 2(10.5) | 1(2.3) |  |  |
|  | (Missing) | 2 (10.0) | 1 (2.2) |  |  |  |  |  |  |
| cDMA | Median (IQR) | 0.0 (-0.1 to 0.0) | 0.0 (0.0 to 0.0) | 0.386 |  | 0.0 (0.0 to 0.0) | 0.0 (0.0 to 0.0) | 0.611 |  |

CPSP, Central post-stroke pain ; NPSS, Non-pain sensory stroke; CDT, cold detection threshold; WDT, warm detection threshold, TSL, thermal sensory limen, CPT, cold pain threshold, HPT, heat pain threshold, PPT, pressure pain threshold, MPT, mechanical pain threshold, MPS, mechanical pain sensitivity, WUR, wind up ratio, MDT, mechanical detection threshold, VDT, vibration detection threshold, PHS, paradoxical heat sensations(0-3), DMA, dynamic mechanical allodynia. QST on the contralesional side is indicated with "c" (e.g., cCDT).
Explorative analysis of QST parameters on the contralesional side using Mann-Whitney U test in the acute phase (3a) and chronic phase (3b). Blue colour means "relative loss of function" compared to DFNS reference values; orange means "relative gain of function" compared to reference values (based on the statistical comparisons given in Tables S4/S5 in the supplement). The p-values given here refer to the statistical comparison (without Bonferroni correction) between CPSP and NPSS. Effect sizes reported are Cliff's d, delta.

**Table 4.** QST parameters in the acute (pre-pain) phase and chronic phase: ipsilesional side.

|  |  | Between group comparison |  |  |  | Between group comparison |  |  |  |
| --- | --- | --- | --- | --- | --- | --- | --- | --- | --- |
| Diagnosis |  | a | Acute |  | d | b | Chronic |  | d |
|  |  | CPSP | NPSS | P |  | CPSP | NPSS | P |  |
| Total N (%) |  | 20 (30.8) | 45 (69.2) |  |  | 20 (30.8) | 45 (69.2) |  |  |
| iCDT | Median (IQR) | -0.2 (-1.2 to 0.4) | 0.1 (-0.9 to 0.9) | 0.328 |  | -0.5 (-1.8 to -0.1) | -0.0 (-1.0 to 0.5) | **0.046\*** | -0.15 |
| iWDT | Median (IQR) | -0.1 (-0.9 to 0.6) | -0.2 (-1.2 to 1.0) | 0.925 |  | -1.1 (-1.7 to -0.4) | -0.3 (-0.9 to 0.6) | **0.036\*** | -0.01 |
| iTSL | Median (IQR) | 0.1 (-0.6 to 0.9) | -0.1 (-0.7 to 0.7) | 0.505 |  | -0.5 (-1.2 to 0.1) | -0.2 (-0.8 to 0.5) | 0.200 |  |
| iCPT | Median (IQR) | 0.2 (-0.7 to 1.3) | -0.1 (-0.7 to 1.0) | 0.696 |  | 0.4 (-0.6 to 1.1) | 0.4 (-0.5 to 1.5) | 0.557 |  |
| iHPT | Median (IQR) | 0.7 (-0.6 to 1.8) | -0.3 (-0.8 to 0.9) | 0.288 |  | -0.7 (-1.2 to -0.2) | -0.0 (-0.9 to 1.3) | **0.018\*** | 0.17 |
| iPPT | Median (IQR) | -0.9 (-1.6 to -0.2) | -0.4 (-1.4 to 0.6) | 0.170 |  | -0.7 (-2.1 to -0.0) | -0.5 (-1.6 to 0.5) | 0.238 |  |
| iMPT | Median (IQR) | 1.2 (0.2 to 2.8) | 1.3 (0.3 to 2.7) | 0.798 |  | 2.0 (0.3 to 3.0) | 1.6 (0.4 to 2.9) | 0.853 |  |
| iMPS | Median (IQR) | 1.4 (0.2 to 1.9) | 1.5 (0.0 to 2.6) | 0.629 |  | 0.9 (0.4 to 2.1) | 1.1 (0.6 to 2.6) | 0.274 |  |
| iWUR | Median (IQR) | -0.7 (-1.0 to 0.2) | -0.2 (-0.8 to 0.7) | 0.319 |  | -0.5 (-0.8 to 0.1) | -0.4 (-0.9 to 0.2) | 0.917 |  |
| iMDT | Median (IQR) | -0.8 (-1.4 to -0.3) | -0.4 (-1.5 to 0.6) | 0.422 |  | -0.7 (-1.8 to 0.4) | -0.3 (-1.7 to 0.6) | 0.491 |  |
| iVDT | Median (IQR) | -0.0 (-0.7 to 0.8) | 0.8 (0.1 to 0.9) | **0.039\*** | -0.32 | 0.1 (-1.0 to 0.7) | 0.8 (0.2 to 1.1) | **0.020\*** | -0.32 |
| iPHS | 0 | 19 (95.0) | 40 (88.9) | 1.000 |  | 17(89.5) | 43(100.0) | 0.090 |  |
|  | 1 | 1 (5.0) | 3 (6.7) |  |  | 1(5.3) | 0(0.0) |  |  |
|  | 2 | 0 (0.0) | 1 (2.2) |  |  | 0(0.0) | 0(0.0) |  |  |
|  | 3 | 0 (0.0) | 0 (0.0) |  |  | 1(5.3) | 0(0.0) |  |  |
|  | (Missing) | 0 (0.0) | 1 (2.2) |  |  |  |  |  |  |
| iDMA | Median (IQR) | 0.0 (0.0 to 0.0) | 0.0 (0.0 to 0.0) | 0.547 |  | 0.0 (0.0 to 0.0) | 0.0 (0.0 to 0.0) | 0.382 |  |

CPSP, Central post-stroke pain ; NPSS, Non pain sensory stroke; QST, Quantitative Sensory Testing; CDT, cold detection threshold; WDT, warm detection threshold; TSL, thermal sensory limen; CPT , cold pain threshold; HPT, heat pain threshold; PPT, pressure pain threshold; MPT, mechanical pain threshold; MPS mechanical pain sensitivity; WUR, wind up ratio; MDT, mechanical detection threshold; VDT, vibration detection threshold; PHS, paradoxical heat sensations(0-3); DMA, dynamic mechanical allodynia. QST on the ipsilesional side is indicated with “i” (e.g., iCDT).
Explorative analysis of QST parameters on the ipsilesional side using Mann-Whitney U tests. Blue colour means "relative loss of function" compared to DFNS reference values; orange means "relative gain of function" compared to reference values (based on the statistical comparisons given in Tables S4/S5 in the supplement). The p-values given here refer to the statistical comparison (without Bonferroni correction) between CPSP and NPSS. Effect sizes reported are Cliff's d, delta.

**Table 5.** QST parameters in the acute (pre-pain) phase and chronic phase: side-to-side differences.

| Diagnosis |  | Between group comparison |  |  |  | Between group comparison |  |  |  |
| --- | --- | --- | --- | --- | --- | --- | --- | --- | --- |
|  |  | a. | Acute |  | d | b. | Chronic |  | d |
|  |  | CPSP | NPSS | P |  | CPSP | NPSS | P |  |
| Total N (%) |  | 20 (30.8) | 45 (69.2) |  |  | 20 (30.8) | 45 (69.2) |  |  |
| sdCDT | Median (IQR) | -1.1 (-2.8 to -0.1) | -0.8 (-1.4 to 0.7) | 0.118 |  | -0.6 (-1.4 to 0.6) | -0.0 (-1.0 to 0.6) | 0.388 |  |
| sdWDT | Median (IQR) | -1.5 (-3.0 to -0.5) | -0.8 (-2.1 to 0.4) | 0.121 |  | -0.6 (-2.0 to 0.1) | -0.6 (-1.3 to 0.3) | 0.459 |  |
| sdTSL | Median (IQR) | -1.9 (-3.2 to -0.5) | -0.8 (-1.7 to 0.2) | **0.030\*** | -0.34 | -1.0 (-2.4 to 0.6) | -0.6 (-1.6 to 0.4) | 0.609 |  |
| sdCPT | Median (IQR) | -0.4 (-1.1 to 0.2) | 1.2 (0.0 to 2.3) | **<0.001\*** | -0.55 | -0.1 (-0.7 to 0.7) | 0.3 (-0.1 to 1.6) | 0.075 |  |
| sdHPT | Median (IQR) | -2.0 (-3.1 to -0.2) | -0.1 (-0.9 to 0.5) | **0.012\*** | -0.40 | -0.4 (-1.0 to 0.3) | 0.0 (-0.9 to 0.8) | 0.631 |  |
| sdPPT | Median (IQR) | -0.6 (-1.5 to 0.6) | 0.0 (-0.6 to 0.6) | 0.112 |  | -0.2 (-1.4 to 0.5) | 0.1 (-0.8 to 0.7) | 0.241 |  |
| sdMPT | Median (IQR) | -0.6 (-2.1 to 0.1) | 0.0 (-1.2 to 1.9) | 0.090 |  | -0.1 (-2.4 to 1.3) | 0.0 (-0.9 to 0.9) | 0.467 |  |
| sdMPS | Median (IQR) | -1.0 (-3.1 to 0.3) | -0.5 (-1.3 to 0.2) | 0.280 |  | -0.2 (-0.9 to 0.7) | -0.1 (-0.4 to 0.7) | 0.486 |  |
| sdWUR | Median (IQR) | 0.6 (-0.1 to 1.7) | 0.2 (-0.6 to 1.0) | 0.056 |  | 0.3 (0.1 to 0.6) | 0.2 (-0.2 to 0.6) | 0.330 |  |
| sdMDT | Median (IQR) | -1.3 (-3.6 to 1.4) | -0.7 (-2.3 to 0.3) | 0.842 |  | -1.6 (-1.9 to -0.8) | 0.3 (-1.8 to 1.4) | **0.025\*** | -0.03 |
| sdVDT | Median (IQR) | -0.1 (-1.3 to 0.0) | 0.0 (0.0 to 0.0) | 0.068 |  | 0.0 (-1.1 to 0.0) | 0.0 (0.0 to 0.0) | 0.348 |  |

CPSP, Central post-stroke pain ; NPSS, Non pain sensory stroke; QST, Quantitative Sensory Testing; CDT, cold detection threshold; WDT, warm detection threshold; TSL, thermal sensory limen; CPT , cold pain threshold; HPT, heat pain threshold; PPT, pressure pain threshold; MPT, mechanical pain threshold; MPS mechanical pain sensitivity; WUR, wind up ratio; MDT, mechanical detection threshold; VDT, vibration detection threshold; PHS, paradoxical heat sensations(0-3); DMA, dynamic mechanical allodynia. QST side-to-side differences are indicated with “sd” (e.g., sdCDT).
Explorative analysis of QST parameters for side-to-side differences using Mann-Whitney U tests. Blue colour means "relative loss of function" compared to DFNS reference values; orange means "relative gain of function" compared to DFNS reference values (based on the statistical comparisons given in Tables S4/S5 in the supplement). The p-values given here refer to the statistical comparison (without Bonferroni correction) between CPSP and NPSS. Effect sizes reported are Cliff's d, delta.

### QST in the chronic phase after stroke

The QST exams in the chronic post-stroke phase were performed between 39 - 361 days (mean *=* 198.4, SD = 48.0) after the onset of stroke.

- **Contralesionally**, CPSP patients continued to show significant thermal hypoesthesia (_c_CDT *U*=585, *P*=0.007; _c_WDT *U*=569, *P*=0.014) and hypoalgesia to cold and heat stimuli (_c_CPT *U*=551, *P*=0.030; _c_HPT *U=*539, *P*=0.046) compared to NPSS patients. Further, CPSP patients had more pronounced mechanical hypoesthesia (cMDT *U*=590,*P*=0.047) and pallhypoesthesia (cVDT *U=*617, *P*=0.010) compared to NPSS patients.
- **Ipsilesionally**, similar patterns to the acute phase of thermal hypoesthesia (_i_CDT *U*=539, *P*=0.046, _i_WDT *U*=546, *P*=0.036), and heat hypoalgesia (_i_HPT *U=*564, *P*=0.018) were observed in CPSP patients. Further, significant disparities in pallesthesia when compared to NPSS patients (_i_VDT *U*=600, *P*=0.020) were observed.
- **Side-to-side comparisons** highlighted mechanical hypoesthesia *U*=608, P*=*0.025 (uncorrected) predominantly on the contralesional side in CPSP patients.

Results are detailed in Table 3-5b and supplementary Figure S3A-C. Combinations of sensory abnormalities, as previously suggested^37^ are detailed in the supplementary results and Table S6 and S7.

### Longitudinal analysis of QST parameters

No significant group and time interactions were seen on the contralesional side (supplementary Table S8). Significant interactions were seen ipsilesionally in the _i_WDT (*F* (1,52)=5.0, *P*=0.029), and _i_HPT (*F* (1,48)=6.5, *P=*0.014, *P*=0.042 after Bonferroni correction) parameters. Post-hoc tests using bootstrapping revealed that only the interaction for _i_HPT remained significant *P=*0.004. This was likely driven by the CPSP group, which showed a significant loss of function in ipsilesional pain perception between acute and chronic phases _i_HPT *t* (19)=3.31, *P*=0.004 (uncorrected) which was not evident in NPSS patients. This indicates that a progressive loss of ipsilesional heat pain perception is specific for CPSP.

On the side-to-side differences, group and time interactions were significant for _sd_HPT (*F* (1,40)=4.2, *P*=0.046). Post-hoc tests with bootstrapping also revealed that the group and time interaction for _sd_HPT was significant *P*=0.002. This was driven by the CPSP group, which showed a smaller side-to-side difference in chronic heat pain perception compared to acute *t* (19)=-3.15, *P*=0.005 (uncorrected).

### Bilateral QST abnormalities

Both CPSP and NPSS groups show significant deviations from the DFNS reference values see supplementary table S4 and S5. In the acute setting CPSP patients had a bilateral loss of function in pressure pain, while NPSS patients had a bilateral gain of function in mechanical pain. In the chronic setting CPSP patients had a bilateral loss of function in temperature perception and mechanical detection and a gain of function in mechanical pain. NPSS patients had bilateral gain of function in cold pain and mechanical pain.

### Predictor analysis

Univariate binary logistic regression was conducted to assess the influence of various factors on the development of CPSP, including sex, age, neurological impairment, sleep quality, QoL, depression, and QST parameters acutely post-stroke. The variables found to be significant predictors for later development of CPSP are displayed in supplementary table S9.

Subsequently, a multiple logistic regression analysis was performed, incorporating significant parameters from the previous binary logistic regression. The analysis revealed that hypersensitivity to blunt pressure (_c_PPT: OR 0.23, 95% CI 0.04-0.70, *P*=0.034) was a significant predictor. The Nagelkerke’s R² for the multiple logistic regression was 68.7%, indicating a good fit of the model.

To further streamline the model, a reduced analysis was conducted, including NIHSS, sex, _c_CDT, _c_PPT, _sd_CPT, _sd_HPT, and _c_VDT. This led to an improved AIC from 60.5 to 51.5 indicating a better fit of the reduced model, though the c-statistic reduced slightly suggesting that the full model had better discrimination. Nagelkerke’s R^2^ also reduced slightly from 68.7% to 65%. The results indicated that _c_PPT (OR 0.24, 95% CI 0.07-0.56, *P*=0.006), and _sd_CPT (OR 0.54, 95% CI 0.28-0.89, *P*=0.029) remained significant predictors (Figure 3C).

**Figure 3.**
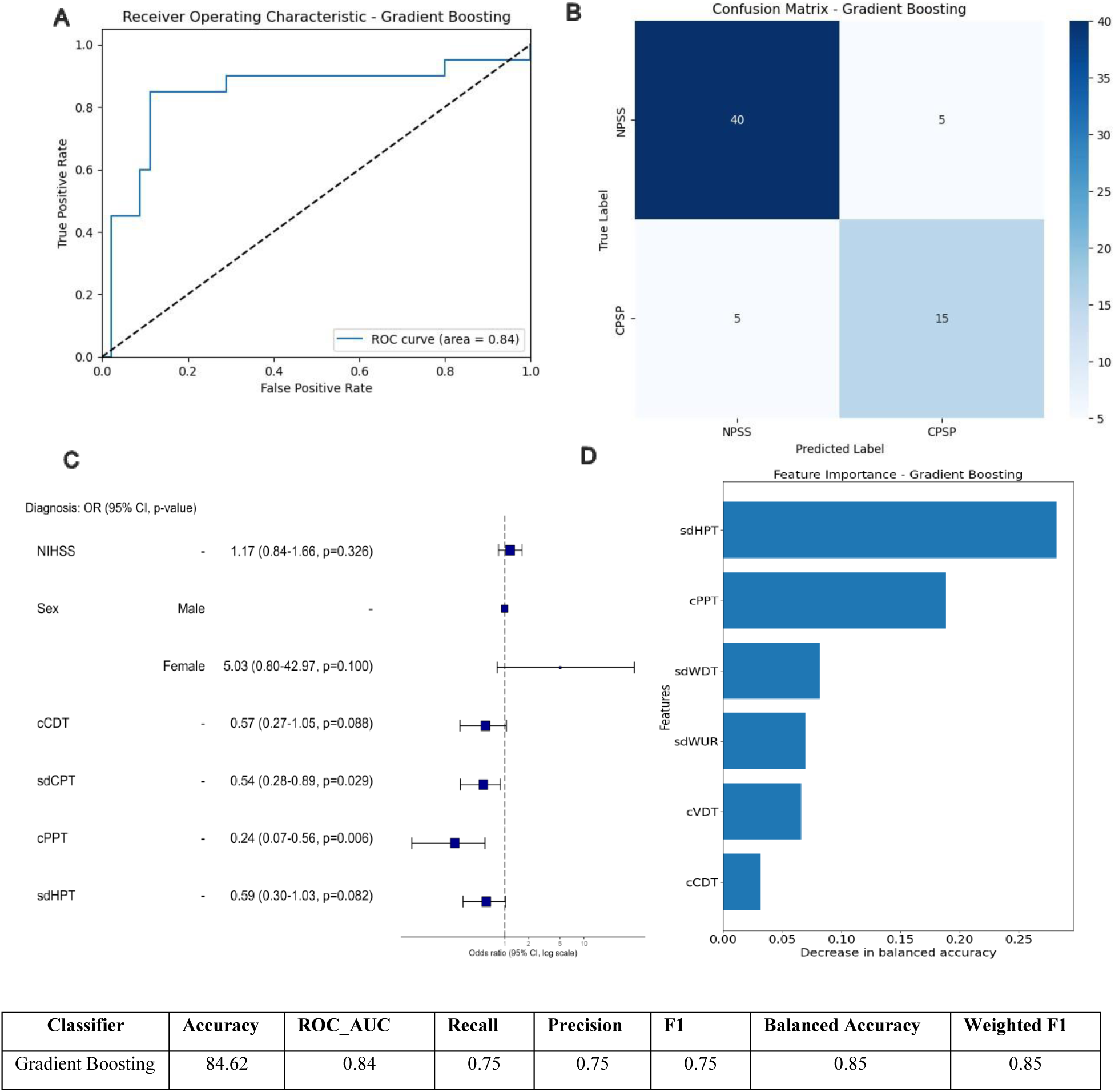
Prediction of pain occurrence from the acute setting QST parameters. CPSP, Central post-stroke pain; QST, Quantitative Sensory Testing; CDT, cold detection threshold; CPT, cold pain threshold; PPT, pressure pain threshold; HPT, heat pain threshold; NIHSS, national institutes of health stroke scale; QST on the contralesional side is indicated with “c” (e.g., cCDT). Side-to-side differences between the ipsilesional and the contralesional side are indicated with “sd” (e.g., sdCDT). Gradient boosting classification (with leave-one out cross validation) based on Quantitative Sensory Testing (QST) findings in acute stage to predict pain occurrence. (A) ROC curve of the gradient boosting classifier. (B) Confusion matrix. (C) Odds ratio plot of the logistic regression. (D) Feature importance showing the contribution of the 6 QST features included in the classifier model.

### Classification of CPSP and NPSS patients before pain onset

We used a Gradient Boosting classifier with LOOCV to categorise CPSP (*n*=20) vs. NPSS (*n=*45) patients before the onset of pain in the acute phase (Figure 3). The model correctly classified 15 out of 20 CPSP patients, giving an 85% accuracy and an AUC-ROC of 0.84. Both precision and recall were 0.75, indicating balanced classification capabilities, further supported by a consistent F1-score of 0.75. Feature importance identified the QST parameters _sd_HPT and _c_PPT, aligning with statistical differences observed between the groups.

## Discussion

### Main findings

With the aim of identifying potential predictors of CPSP, we present the results of the first prospective and longitudinal evaluation of quantitative sensory testing (QST) profiles in patients with acute somatosensory stroke. Of the 75 study participants, 26 (34.7%) developed CPSP within eight months. Prior to the onset of pain, CPSP patients compared to NPSS patients had more severe cold hypoesthesia confirming one of our two hypotheses. The additional exploratory analysis showed that almost all thermal QST parameters differed between CPSP and NPSS patients on the symptomatic side or in the side-to-side difference before pain onset in univariate comparisons, with one parameter, the side-to-side difference of cold pain perception, being significant after Bonferroni correction. Using a gradient-boosting approach we were able to correctly classify future CPSP patients, before they developed pain, with an overall accuracy of 0.85, a recall of 0.75, and a precision of 0.75. Another notable finding is that approximately 80% of patients with unilateral somatosensory stroke (CPSP and NPSS) had bilateral sensory QST changes (> 2 SD from reference values) in both the acute and chronic phases.

### CPSP after Somatosensory Stroke

In our cohort, 34.7% of stroke patients developed CPSP, a rate higher than that reported in previous somatosensory stroke studies.^21^ Potentially, this could arise due to a higher drop out of NPSS patients. The time delay between stroke onset and onset of pain was between 1 week and 8 months (Table 1) with most patients developing pain within 1 month, which agrees with previous observations.^38^ The age and sex distribution of all included patients with somatosensory stroke (65 (55.5-70) years, f:m 1:1.5) were within the expected range at our institution.^39^ In our study, female sex as well as the severity of stroke were associated with higher risk of CPSP. A study by Hansen *et al.*^2^ has also reported that patients with CPSP were more often female and younger, however other studies have reported no sex differences.^2^

### QST in acute/subacute stroke

While prior research has examined acute stroke patients, QST had not been applied in these investigations.^11^ We found substantial differences between CPSP and NPSS patients in the acute/subacute “pre-pain” stroke phase specifically, we found compelling evidence for, an increased cold detection threshold (CDT) in CPSP patients.

Furthermore, in our exploratory analyses we found that CPSP patients mainly differed from NPSS patients in terms of thermal and nociceptive parameters, which further supports previous interpretations that CPSP is primarily a deficit of the spinothalamic tract, rather than a defici of dorsal column function.^40^ When comparing the acute QST z-values from our cohort with the reference values of the DFNS cohort, CPSP patients showed a significant loss of function in temperature and mechanical perception, and hypoalgesia to pressure pain. NPSS patients, on the other hand, showed a loss of function to temperature and mechanical perception and pronounced hyperalgesia to cold and mechanical pain.

Overall, loss of function occurred much more frequently in CPSP patients and gain of function more frequently in NPSS patients. For example, only in the latter group did cold painperception (_c_CPT, _sd_CPT) and mechanical pain perception and sensitivity (_c_MPT, _c_MPS) deviate from the DFNS reference values as gain of function. This indicates a tendency for hyperalgesia to cold and mechanical stimuli on the contralesional side.

### Towards pain-prediction

The exploratory findings that almost all temperature based QST measures differed between CPSP and NPSS patients, before the pain developed and one of them _sd_CPT remained significant after Bonferroni correction, provided further evidence that prediction of pain based solely on sensory findings might be possible.

Indeed, the gradient-boosting approach to predicting CPSP resulted in a good overall accuracy of 0.85. Overall, we believe that these results represent an important step towards clinically meaningful pain prediction. As a next step, we call for a confirmatory multicentre study, the results of which could be the basis for future clinical trials on pain prevention in CPSP patients. Such a trial, might also consider results of a recent study which, based on data from the UK Biobank, has shown a set of biopsychosocial factors including sleep, neuroticism, mood, life stressors, and body mass index to be sufficient to predict the development and spread of chronic pain.^41^

### Bilateral sensory deviations

Both patient groups showed perception deficits not only on the contralesional side but also on the ipsilesional side, compared to the reference values from the DFNS, both acutely and at the last QST follow-up. The ipsilesional side is often labelled as “unaffected” by the stroke and serves as a sensory control area in many studies.^11,19^ However, we and others have previously shown that bilateral perception deficits occur in the chronic stage post stroke.^18,42–44^ These bilateral sensory symptoms were not limited to subjects with medulla or paramedian pons lesions, where bilateral facial and perioral symptoms are known to occur, but were observed also on the extremities. In a case series of six patients with CPSP, patients developed symptoms in contralateral (i.e. ipsilesional) counterparts of the body areas where initial pain was most severe.^44^ To our knowledge, ipsilesional sensory symptoms have never been explored in the acute stage post stroke.

### Pathophysiological implications

Three aspects of the QST differences between CPSP and NPSS patients in the acute and chronic phases of stroke are striking. (i) Most marked differences appear to occur for temperature-related parameters, particularly in relation to cold perception. (ii) For almost all sensory parameters, CPSP patients show a relative loss of function compared to NPSS patients in the acute and in the chronic stage. (iii) Both groups exhibit bilateral changes with the ipsilesional side developing QST abnormalities overtime, specific to the CPSP group.

The involvement of the spinothalamic tract has been suggested to be a necessary but not sufficient condition for CPSP.^7^ Lesion studies have shown that thalamic lesions in pain patients often affect the VPL nucleus of the thalamus where the STT is thought to terminate,^5,7^ as well as the anterior nucleus pulvinaris.^6^ Furthermore, a case report has shown that delayed onset of CPSP after thalamic haemorrhage could be due to perilesional neural degeneration of the STT.^45^ The authors used diffusion tensor imaging that showed progressive thinning and tearing of the STT with progression of the patient’s pain symptoms.^45^ Also, in our CPSP cohort spinothalamic tract functions were predominantly affected.

In our study, some CPSP and NPSS patients exhibit mechanical hypoesthesia, which indicates an affection of the lemniscal pathway or its projections.^12^ As previously reported an affection of the medial lemniscus pathway is not necessarily associated with CPSP.^12,19^

In our patient cohort most CPSP patients mainly display “sensory loss” one of the proposed phenotype clusters of peripheral neuropathic pain proposed by Baron et al.^46^ The loss of function in CPSP patients is likely the correlate of central deafferentation due to neuronal damage.^47^ These QST findings “before pain” align with results from other QST models in various types of neuropathic pain, primarily of peripheral origin, where patients’ chronic pain is manifested.^46^ Additionally, they correspond with human surrogate models of functional transient denervation, demonstrating that A-fibre compression block results in a significant reduction in thermal and mechanical detection, accompanied by paradoxical heat sensations.^48^ Others have also termed this painful hypoesthesia,^40,46^ with spontaneous pain arising due to ectopic activity generated by deafferented central nociceptive neurons. Interestingly, NPSS patients’ QST parameters were more in line with a pattern of thermal or mechanical hyperalgesia, though these patients did not develop pain during our follow up period. This raises the question of whether increased sensitivity to thermal pain, combined witha mildly affected thermal discrimination ability, could be indicative of a mechanism which protects against CPSP. This is suggested by our regression analysis, showing that a hyperalgesia to cold, and blunt pressure on the contralesional side, decreased the risk of developing CPSP.

While the pathophysiology of bilateral sensory abnormalities in CPSP is not fully understood, a possible explanation includes indirect sensory pathways that do not decussate in the spinal cord.^44,49^ Furthermore, bilateral cortical atrophy within the somatosensory cortex, insula, and prefrontal cortex has been described in the chronic stage of CPSP.^50^ These areas are involved in the sensory-discriminative as well as emotional-affective aspects of pain perception.^20,42,50,51^ As the CPSP pathology progresses over time, ipsilesional symptoms could also be related to disinhibition mechanisms due to damage to central inhibition pathways or to central sensitisation due to ectopic activity.^52^

Dynamic central disinhibition of descendent inhibitory pathways has been proposed^53^ and could affect the perception of bilateral peripheral stimuli over time. Furthermore, impaired functional connectivity between cortical and subcortical structures has been described in the context of peripheral pain syndromes^54^ and similar processes could occur after a stroke. CPSP can also be regarded as a phenomenon involving both peripheral and central sensitisation processes, a common concept in the context of peripheral neuropathic pain conditions.^55^ It has been hypothesized that a stroke lesion causes central sensitisation of somatosensory neurons in response to peripheral sensory stimulation. A small study on eight CPSP patients supported this hypothesis by showing that a peripheral lidocaine nerve block abolished both, perception of peripheral input as measured by QST and pain.^56^ Despite having excluded patients with peripheral polyneuropathy it is in principle possible that subclinical involvement of the peripheral nervous systems due to, concomitant diseases including diabetes mellitus, arterial hypertension and adiposity can explain bilateral QST findings.^57^

### Limitations

Our study’s limitations include reliance on DFNS reference data instead of a local control group, potentially overlooking centre and examiner variability. Although QST examiners were DFNS-trained, their awareness of participants’ pain status could introduce bias. Furthermore, it is possible that stroke patients differ *a priori* from healthy individuals. Our strict inclusion and exclusion criteria mean that our study population is carefully selected and well defined, but this also leads to a relatively small and unbalanced sample size. However, this reflects the reality in clinical practice as most patients with somatosensory stroke do not develop pain. The observation time was limited to eight months, as CPSP mainly occurs within the first few months after a stroke.^1,2,21^ However, a later onset of CPSP in the NPSS patients cannot be ruled out.^13,38^ Lastly, the risk of overfitting in our classification model due to the small sample size necessitates cautious interpretation and validation in larger studies.

## Conclusions

Our study is pioneering in prospectively applying QST to acute somatosensory stroke patients and identifying early sensory differences predictive of CPSP development. Notably, differences in cold perception between CPSP and NPSS patients shortly after stroke were found to be significant. Early classification of impending CPSP based on QST-patterns seems possible. Both NPSS and CPSP patients showed bilateral sensory changes in the acute and chronic post stroke stages. The early post-stroke phase is critical, highlighting the importance of timely interventions. Integrating clinical assessments with QST can enhance the identification of patients prone to central neuropathic pain, enabling early therapeutic measures to mitigate pain chronification.

## Supporting information

Supplementary methods and results

## Data Availability

All data produced in the present study are available upon reasonable request to the authors

## Acknowledgements

We would like to extend many thanks to all colleagues from the Stroke Imaging Team and the Trial Team of the Centre for Stroke Research Berlin (CSB) and from the Department of Neurology for their contribution to patient recruitment, study performance, and data acquisition. We would like to thank Dr. Nico Scherf for his valuable comments on the machine learning classification analysis. The authors acknowledge that ChatGPT and DeepL were used to edit grammar and improve the syntax of the paper. Furthermore, the authors thank Joshua Grant for proofreading our final manuscript. Finally, the authors thank all the patients who participated in the study.

## Author contributions

Conception and design of the study: TK, AV, GJJ Acquisition and analysis of data: SA, EP, JM, KV, EA, XC, TK, SH, AV, GJJ Drafting a significant portion of the manuscript or figures: SA, EA, JM, AV, GJJ

