## Supplementary methods and results for "Prediction of Central Post-Stroke Pain by Quantitative Sensory Testing"

**Supplementary materials and methods**

**Exclusion criteria**

Exclusion criteria comprised the inability to provide informed consent or to undergo MRI, immobility, severe paresis, brain atrophy, leukoencephalopathy, stroke lesions affecting more than 50% of the territory supplied by the middle cerebral artery (MCA), acute stroke lesions in multiple cerebral arterial territories, pre-existing lesions within the somatosensory system, concomitant neurological and malignant diseases or pain due to other conditions indistinguishable from CPSP (e.g., polyneuropathy, other entities of post-stroke pain).

**Clinical sensory testing**

In addition to the QST assessment, patients were examined neurologically by a study neurologist. The assessment included a detailed neurological examination with emphasis on the somatosensory system, assessment of light touch with cotton wool to detect hyperesthesia, hypoesthesia, and allodynia. Thermaesthesia was evaluated using two objects of different temperatures, pallesthesia with a 128 Hz tuning fork, and hypo- or hyperalgesia were evaluated by pin-prick testing. Patients were tested on both sides for increased or decreased sensations of touch, temperature, vibration, and pain. The side corresponding to the lesion was interpreted as “affected” and the other side as “unaffected”. Given the focus on standardised quantitative sensory testing in this study, the results of these clinical exams are provided in the supplement (Table S2).

**QST**

To perform these measurements, alternating heat and cold stimuli (Thermode thermal sensory analyser (Model TSAII 2001 from Medoc Ltd., Israel), von Frey hairs (Optihair2 set, Marstock Nervtest, Germany), pin prick needles (Pinprick, MRC Systems GmbH, Germany), cotton wool, standardised brushes (Somedic, Sweden), a tuning fork (64 Hz Rydel-Seiffer), and a pressure algometer (FDK20, Wagner Instruments, Greenwich, CT, USA) were used.^1^

To achieve normal distribution the parameters CDT, WDT, TSL, MDT, MPT, MPS, WUR, PPT, and DMA were logarithmically transformed.^2^ Raw data of CPT, HPT, VDT, and logarithmic data were then z-transformed. PHS and positive DMA values were not z-transformed and given as raw values since they usually do not occur in a healthy population. Z-values above "0" indicate a “gain of function” (e.g., hyperesthesia/hyperalgesia to cold, warm, temperature alterations, cold pain, heat pain, mechanical stimulation, mechanical pain, wind-up phenomenon, pressure pain) while negative z-values are considered as a “loss of function” (hypoesthesia/hypoalgesia). If individual z-values were outside the 95% CI (z-score >1.96 or <-1.96) of the reference group, the values were designated as “pathological hyperesthesia "gain of function" or hypoesthesia "loss of function".^1,2^ The confidence interval for the CPSP and NPSS groups was adjusted for the numbers of observations by dividing the standard confidence interval through the root of the number of observations ((±1.96/ √ (number of observations) for two-sided confidence intervals and 1.64/√ (number of observations) for one-sided confidence intervals (VDT). The number of observations was *n =* 26 for CPSP and *n =* 49 for NPSS, indicating a deviation from the reference values (for the respective group) if the mean QST values lie outside the confidence interval of ±0.38 two-sided, or ±0.32 one-sided for CPSP patients and outside the confidence interval ±0.29 two-sided, or ±0.23 one-sided for NPSS patients. As suggested by Magerl *et al.*^3^, a non-paired t test for data that have been z-normalised was used to compare our patient cohort with the reference data.^3^ The equation is as follows and the distribution of z-values of the control group is always given as mean *=* 0 and SD = 1.

*t = (mean_exp_ – mean_con_) / square root (SD^2^/n_exp_ + SD^2^/n_con_)* (1)

mean_co_*_n =_* 0	(2)

SD_co_*_n =_* 1	(3)

n_Exp_ = n_Con_	(4)

QST findings were categorised based on sensory patterns corresponding to either small or large fibre dysfunction, following the protocol reported by Maier *et al.*^4^ To achieve this, individual z-scores outside the 95% CI were categorised as loss of function (L1-3) or gain of function (G1-3).^4^ Signs of hypoesthesia to thermal stimuli (z-values < -1.96 in _c_CDT, _c_WDT) were designated L1 and signs of hypoesthesia to mechanical stimuli (z-values < -1.96 in _c_MDT, _c_VDT) were designated L2. Signs of hyperalgesia to thermal stimuli (z-values > 1.96 in _c_CPT, _c_HPT) were designated G1 and signs of hyperalgesia to mechanical stimuli (z-values > 1.96 in _c_MPT, _c_MPS, _c_PPT or positive _c_DMA values) were designated G2. If abnormal z-values indicating either loss or gain of function to both thermal and mechanical stimuli occurred, they were designated L3 or G3, respectively.^4^

**Supplementary results**

**Acute phase reference group comparison (Supplementary tables S4, S5)**

**Contralesional**

When comparing CPSP patients to the reference group they showed primarily a loss of function in thermal perception, pressure pain, and mechanical detection. NPSS patients showed loss and gain of function in the acute setting. The NPSS group showed loss of function in the same thermal and mechanical detection parameters, excluding pressure pain as the CPSP group and gain of function in cold pain and mechanical pain parameters. These data including the statistical findings are given in Table S4 and S5 in the supplement. Parameters that significantly differ from the reference DFNS values are highlighted in blue (loss of function) and orange (gain of function).

**Ipsilesional**

In comparison to the reference group, CPSP patients showed a loss of function in pressure pain. They also exhibited a gain of function in mechanical pain threshold and sensitivity. NPSS patients showed similar patterns on the ipsilesional side with a gain of function in mechanical pain threshold and sensitivity.

**Side to side differences**

When comparing CPSP and NPSS to the reference collective, most side differences showed an abnormal deviation on the contralesional side. The contralesional side primarily showed a loss of function compared to the ipsilesional side for both CPSP and NPSS patients. Only in cold pain perception did NPSS patients have a gain of function compared to the ipsilesional side. CPSP patients had a gain of function in the WUR suggesting the presence of hyperpathia on the contralesional side compared to the ipsilesional one.

**Chronic phase reference group comparison (Supplementary tables S4, S5)**

**Contralesional**

In comparison to the reference group, CPSP patients continued to show a hypoesthesia in thermal parameters _c_CDT, _c_WDT, _c_TSL, in mechanical detection and a hypoalgesia to pressure pain. Furthermore, CPSP patients exhibited hyperalgesia to mechanical pain, which was not present in the acute setting.

In comparison to the reference group NPSS patients also continued to have hypoesthesia to thermal parameters (_c_WDT, _c_TSL) and hyperalgesia to cold pain, and mechanical pain.

**Ipsilesional**

Compared to the reference values, CPSP patients showed ipsilesional losses of function in thermal and mechanical detection, and heat pain, which were not statistically significant in the acute phase, in addition to pressure pain perception. A gain of function is reported in mechanical pain parameters (_i_MPT). In NPSS patients we only saw a gain of function in cold pain and mechanical pain in the chronic setting.

Thus, overall, more parameters than in the acute phase differed significantly (not Bonferroni corrected) between the two groups and deviated from the reference values in the chronic setting (please see the longitudinal ANOVA analyses assessing the changes over time of both groups below).

### **Sensory abnormality combinations (Supplementary tables S6, S7)**

To effectively represent various combinations of sensory abnormalities, a coding system was implemented as previously suggested by Maier *et al.*^4^ and detailed in Supplementary Table S6 and S7.

In the acute phase CPSP patients most frequently displayed a *mixed mechanical and thermal loss of function* (*n*=8;42%) while NPSS patients most frequently displayed *mixed hyperalgesia* to mechanical and thermal stimuli (*n*=16;37%). The second most common combination of abnormalities for both groups was *mechanical only gain of function*.

In the chronic phase CPSP patients most commonly showed *mechanical only gain of function* (*n=*8; 42%) whereas NPSS patients most commonly had no loss of function in thermal or mechanical detection (*n*=18;44%).

1.	Rolke R, Baron R, Maier C, et al. Quantitative sensory testing in the German Research Network on Neuropathic Pain (DFNS): standardized protocol and reference values. *Pain*. Aug 2006;123(3):231-243. doi:10.1016/j.pain.2006.01.041

2.	Rolke R, Magerl W, Campbell KA, Schalber C, Caspari S, Birklein F, Treede RD. Quantitative sensory testing: a comprehensive protocol for clinical trials. *Eur J Pain*. Jan 2006;10(1):77-88. doi:10.1016/j.ejpain.2005.02.003

3.	Magerl W, Krumova EK, Baron R, Tölle T, Treede RD, Maier C. Reference data for quantitative sensory testing (QST): refined stratification for age and a novel method for statistical comparison of group data. *Pain*. Dec 2010;151(3):598-605. doi:10.1016/j.pain.2010.07.026

4.	Maier C, Baron R, Tölle TR, et al. Quantitative sensory testing in the German Research Network on Neuropathic Pain (DFNS): somatosensory abnormalities in 1236 patients with different neuropathic pain syndromes. *Pain*. Sep 2010;150(3):439-450. doi:10.1016/j.pain.2010.05.002
