## Supplementary figures for "Prediction of Central Post-Stroke Pain by Quantitative Sensory Testing"

**Table S1: A. Somatosensory stroke study protocol**

| **Visit** | | **V0** | | **V1** | **V2** | | **V3** | **V4** | | **V5** | **V6** |
| --- | --- | --- | --- | --- | --- | --- | --- | --- | --- | --- | --- |
| Time point | | 0d | | 1-3d | 4-10d | | 30d | 90d | | 180d | 1y |
| Inclusion and exclusion criteria | | x | |  |  | |  |  | |  |  |
| Past medical history, medication history | | x | | x | x | | x | x | | x | x |
| Scores  (NIHSS, mRS, Barthel) | | x (pre) | | x | x | | x | x | | x |  |
| Clinical examination | | x | | x | x | | x | x | | x |  |
| Questionnaires  (GDS-30, SF-12, PSQI, DSF*, PAINdetect*, SES*) | |  | | x | x | | x | x | | x |  |
| QST | |  | | x | x | | x | x | | x |  |
| MRI | |  | | x | x | | x | x | | x |  |
| **B. Average days from stroke and number of patients completing each follow up visit** | | | | | | | | | | | |
| Visit | Average (days) | | Range(days) | | | SD | | | Number of Patients  (n=75) | | |
| V1 | 2 | | 1-3 | | | 1 | | | 41 | | |
| V2 | 6 | | 4-10 | | | 2 | | | 53 | | |
| V3 | 45 | | 22-77 | | | 14 | | | 60 | | |
| V4 | 106 | | 54-144 | | | 16 | | | 67 | | |
| V5 | 207 | | 175-361 | | | 34 | | | 72 | | |
| V6 | 392 | | 235-541 | | | 84 | | | 14 | | |

*Only if pain indicated

NIHSS= National Institutes of Health Stroke Scale, mRS = Modified Rankin scale, PSQI = Pittsburgh Sleep Quality Index, GDS-30 = Geriatric Depression Scale, SF-12-MCS = short form 12 mental component score, SF-12-PCS = short form 12 physical component score, DSF= Deutsche Schmerzfragebogen (German pain questionnaire), SES = Schmerzempfindungsskala (pain perception scale). QST = quantitative sensory testing.

**Table S2: Positive and negative sensory symptoms CPSP vs. NPSS**

| A. | Acute phase | | | | | B. Chronic phase | | | | |
| --- | --- | --- | --- | --- | --- | --- | --- | --- | --- | --- |
|  | CPSP  N=20 (%) |  | NPSS  N=45 (%) |  | p-value | CPSP  N=20 (%) |  | NPSS  N=45 (%) |  | p-value |
| Positive symptoms |  | Missing |  | Missing |  |  | Missing |  | Missing |  |
| Hyperaesthesia | 1(5%) | 4(20%) | 2(4.4%) | 10(22.2%) | 1.000 | 8(40%) | 1(5%) | 8(17.8%) | 0 | **0.036*** |
| Hyperalgesia | 2(10%) | 4(20%) | 3(6.7%) | 10(22.2%) | 0.910 | 8(40%) | 1(5%) | 7(15.6%) | 0 | **0.022*** |
| Paraesthesia | 10(50%) | 1(5%) | 20(44.4%) | 8(17.8%) | 0.437 | 14(70%) | 1(5%) | 21(46.7%) | 0 | **0.027*** |
| Thermhyperaesthesia | 4(20%) | 4(20%) | 2(4.4%) | 10(22.2%) | 0.147 | 7(35%) | 1(5%) | 8(17.8%) | 0 | 0.074 |
| Mechanical Allodynia | 0 | 4(20%) | 0 | 10(22.2%) | - | 5(25%) | 1(5%) | 1(2.2%) | 0 | **0.003*** |
| Thermal Allodynia | 1(5%) | 4(20%) | 0 | 10(22.2%) | 0.441 | 6(30%) | 1(5%) | 1(2.2%) | 0 | **0.001*** |
| Negative symptoms |  |  |  |  |  |  |  |  |  |  |
| Hypaesthesia | 17(85%) | 0 | 29(64.4%) | 8(17.8%) | 0.096 | 15(75%) | 1(5%) | 25(55.6%) | 0 | **0.046*** |
| Hypalgesia | 10(50%) | 2(10%) | 14(31.1%) | 8(17.8%) | 0.394 | 9(45%) | 1(5%) | 10(22.2%) | 0 | **0.047*** |
| Thermhypaesthesia | 10(50%) | 2(10%) | 9(20%) | 9(20%) | 0.063 | 10(50%) | 1(5%) | 12(26.7%) | 0 | **0.032*** |

CPSP: central post-stroke pain; NPSS: non–pain sensory stroke

A. Positive and negative symptoms reported during clinical examination, either in visit 1 or visit 2, in the first 10 days following a stroke. Two-sided Fisher’s exact test performed.

B. Positive and negative symptoms reported during clinical examination during visit 4 or visit 5. Two-sided Fisher’s exact test performed.

**Table S3: Lesion analysis**

| **Diagnosis** |  | **CPSP** | **NPSS** | **Total** | **p** |
| --- | --- | --- | --- | --- | --- |
| Total N (%) |  | 26 (34.7) | 49 (65.3) | 75 |  |
| Brainstem | **Brainstem** | **6 (23.1)** | **5 (10.2)** | **11 (14.7)** | **0.174** |
|  | Other | 20 (76.9) | 44 (89.8) | 64 (85.3) |  |
| Thalamus | **Thalamus** | **13 (50.0)** | **35 (71.4)** | **48 (64.0)** | **0.081** |
|  | Other | 13 (50.0) | 14 (28.6) | 27 (36.0) |  |
| Cortex | **Cortex** | **7 (26.9)** | **8 (16.3)** | **15 (20.0)** | **0.364** |
|  | Other | 19 (73.1) | 41 (83.7) | 60 (80.0) |  |
| Pathways | **Pathways** | **0** | **1 (2.0)** | **1 (1.3)** | **1.000** |
|  | Other | 26 (100.0) | 48 (98.0) | 74 (98.7) |  |

Complementary lesion analysis looking at differences between CPSP and NPSS in the frequency of lesions in the brainstem, thalamus, cortex or pathways. In this analysis each region (brainstem, thalamus, cortex, pathways) was tested individually.

**Table S4: CPSP patient QST parameter comparison with reference collective**

| **A.** |  | **Acute** | | | | **B. Chronic** | | | | |
| --- | --- | --- | --- | --- | --- | --- | --- | --- | --- | --- |
| QST | N | Mean | SD | T Statistic | P Value | N | Mean | SD | T Statistic | P Value |
| cCDT | 20 | -1.846 | 2.001 | -3.691 | **0.001** | 19 | -1.555 | 1.868 | -3.200 | **0.003** |
| cWDT | 20 | -1.791 | 1.625 | -4.198 | **0.000** | 19 | -1.855 | 1.510 | -4.464 | **0.000** |
| cTSL | 20 | -1.563 | 1.724 | -3.506 | **0.001** | 19 | -1.484 | 1.582 | -3.456 | **0.001** |
| cCPT | 20 | -0.027 | 1.186 | -0.077 | 0.939 | 19 | 0.327 | 1.431 | 0.816 | 0.420 |
| cHPT | 19 | -0.571 | 1.565 | -1.341 | 0.189 | 19 | -0.438 | 1.347 | -1.137 | 0.263 |
| cPPT | 20 | -1.584 | 1.421 | -4.076 | **0.000** | 20 | -1.188 | 1.349 | -3.163 | **0.003** |
| cMPT | 20 | 0.710 | 2.138 | 1.346 | 0.187 | 20 | 1.282 | 2.281 | 2.301 | **0.027** |
| cMPS | 20 | 0.162 | 1.842 | 0.346 | 0.731 | 20 | 0.883 | 1.804 | 1.916 | 0.063 |
| cWUR | 17 | 0.117 | 1.133 | 0.319 | 0.752 | 20 | 0.185 | 1.258 | 0.515 | 0.610 |
| cMDT | 20 | -2.377 | 4.891 | -2.129 | **0.040** | 20 | -1.346 | 1.612 | -3.173 | **0.003** |
| cVDT | 20 | -0.801 | 2.113 | -1.532 | 0.134 | 20 | -0.336 | 1.152 | -0.985 | 0.331 |
| iCDT | 20 | -0.499 | 1.259 | -1.388 | 0.174 | 19 | -0.956 | 1.470 | -2.345 | **0.025** |
| iWDT | 20 | -0.183 | 1.483 | -0.457 | 0.650 | 19 | -1.140 | 1.407 | -2.877 | **0.007** |
| iTSL | 20 | -0.054 | 1.281 | -0.150 | 0.882 | 19 | -0.736 | 1.232 | -2.023 | 0.051 |
| iCPT | 20 | 0.375 | 1.164 | 1.092 | 0.282 | 19 | 0.355 | 1.064 | 1.060 | 0.297 |
| iHPT | 19 | 0.681 | 1.676 | 1.521 | 0.138 | 19 | -0.653 | 0.860 | -2.158 | **0.038** |
| iPPT | 20 | -0.952 | 1.178 | -2.755 | **0.009** | 20 | -0.906 | 1.307 | -2.462 | **0.019** |
| iMPT | 20 | 1.466 | 1.622 | 3.441 | **0.001** | 20 | 1.689 | 1.825 | 3.629 | **0.001** |
| iMPS | 20 | 0.994 | 1.600 | 2.356 | **0.024** | 20 | 0.940 | 1.883 | 1.971 | 0.056 |
| iWUR | 20 | -0.291 | 1.029 | -0.908 | 0.370 | 19 | -0.337 | 0.756 | -1.171 | 0.250 |
| iMDT | 20 | -1.284 | 3.068 | -1.780 | 0.084 | 20 | -0.910 | 1.691 | -2.073 | **0.045** |
| iVDT | 20 | -0.078 | 1.086 | -0.237 | 0.814 | 20 | -0.119 | 1.004 | -0.375 | 0.710 |
| sdCDT | 20 | -1.723 | 2.522 | -2.840 | **0.007** | 19 | -0.796 | 2.304 | -1.382 | 0.176 |
| sdWDT | 20 | -1.871 | 2.019 | -3.714 | **0.001** | 19 | -0.893 | 1.751 | -1.929 | 0.062 |
| sdTSL | 20 | -2.263 | 2.359 | -3.950 | **0.000** | 19 | -1.060 | 2.288 | -1.851 | 0.073 |
| sdCPT | 20 | -0.683 | 1.707 | -1.543 | 0.132 | 19 | -0.032 | 1.440 | -0.080 | 0.937 |
| sdHPT | 19 | -1.507 | 2.418 | -2.510 | **0.017** | 19 | 0.122 | 1.863 | 0.252 | 0.803 |
| sdPPT | 20 | -0.764 | 2.086 | -1.476 | 0.149 | 20 | -0.418 | 1.482 | -1.047 | 0.302 |
| sdMPT | 20 | -1.195 | 2.530 | -1.964 | 0.057 | 20 | -0.709 | 3.228 | -0.938 | 0.354 |
| sdMPS | 20 | -1.938 | 3.390 | -2.452 | **0.019** | 20 | -0.131 | 2.007 | -0.261 | 0.795 |
| sdWUR | 17 | 0.840 | 1.110 | 2.319 | **0.027** | 19 | 0.631 | 1.078 | 1.869 | 0.070 |
| sdMDT | 20 | -1.032 | 3.554 | -1.250 | 0.219 | 20 | -1.046 | 2.291 | -1.871 | 0.069 |
| sdVDT | 20 | -0.915 | 1.576 | -2.192 | **0.035** | 20 | -0.333 | 0.964 | -1.072 | 0.291 |

CPSP: central post-stroke pain; NPSS: non–pain sensory stroke; QST; Quantitative Sensory Testing; CDT, cold detection threshold; CPT, cold pain threshold; HPT, heat pain threshold; MDT, mechanical detection threshold; MPS, mechanical pain sensitivity; MPT ,mechanical pain threshold; PPT, pressure pain threshold; TSL, temperature sensory limen; VDT, vibration detection threshold; WDT, warm detection threshold; WUR, wind- up ratio; QST parameters on the contralesional side are indicated with “c” (e.g., cCDT), on the ipsilesional with “i” (e.g., iCDT). Side-to-side differences are indicated with “sd” (e.g., sdCDT).

Statistical comparisons of QST values to DFNS reference values. A total of 20 CPSP patients were included the N column indicates the number of participants for each QST variable. P-values were obtained by comparing the CPSP group’s Z-standardised QST mean with a virtual reference population mean as suggested by Magerl et al., 2010 assuming a mean of 0 and SD = 1. Alpha level was 0.05, p-values are not Bonferroni-corrected. Blue boxes indicate a loss of function and orange boxes a gain of function.

**Table S5: NPSS patient QST parameter comparison with reference collective**

| **A.** |  | **Acute** | | | | **B. Chronic** | | | | |
| --- | --- | --- | --- | --- | --- | --- | --- | --- | --- | --- |
| QST | N | Mean | SD | T Statistic | P Value | N | Mean | SD | T Statistic | P Value |
| cCDT | 45 | -0.534 | 1.060 | -2.458 | **0.016** | 43 | -0.286 | 1.013 | -1.318 | 0.191 |
| cWDT | 45 | -0.987 | 1.359 | -3.924 | **0.000** | 43 | -0.767 | 1.414 | -2.904 | **0.005** |
| cTSL | 45 | -0.595 | 1.024 | -2.790 | **0.006** | 43 | -0.601 | 1.101 | -2.651 | **0.010** |
| cCPT | 45 | 0.983 | 1.388 | 3.857 | **0.000** | 43 | 0.996 | 1.303 | 3.976 | **0.000** |
| cHPT | 45 | 0.079 | 1.560 | 0.288 | 0.774 | 43 | 0.535 | 1.803 | 1.702 | 0.093 |
| cPPT | 42 | -0.375 | 1.336 | -1.458 | 0.149 | 45 | -0.420 | 1.384 | -1.652 | 0.102 |
| cMPT | 45 | 1.295 | 1.638 | 4.527 | **0.000** | 45 | 1.695 | 1.466 | 6.410 | **0.000** |
| cMPS | 45 | 1.130 | 1.819 | 3.652 | **0.000** | 45 | 1.525 | 1.519 | 5.626 | **0.000** |
| cWUR | 42 | -0.011 | 0.990 | -0.053 | 0.958 | 45 | -0.090 | 0.848 | -0.459 | 0.647 |
| cMDT | 45 | -1.198 | 1.843 | -3.834 | **0.000** | 45 | -0.455 | 1.533 | -1.669 | 0.099 |
| cVDT | 45 | 0.067 | 1.299 | 0.274 | 0.785 | 44 | 0.150 | 1.776 | 0.487 | 0.628 |
| iCDT | 44 | -0.154 | 1.239 | -0.642 | 0.523 | 43 | -0.157 | 1.005 | -0.726 | 0.470 |
| iWDT | 44 | -0.184 | 1.296 | -0.745 | 0.459 | 43 | -0.324 | 1.346 | -1.265 | 0.209 |
| iTSL | 44 | -0.090 | 0.957 | -0.433 | 0.666 | 43 | -0.265 | 0.923 | -1.276 | 0.205 |
| iCPT | 44 | 0.258 | 1.234 | 1.076 | 0.285 | 43 | 0.559 | 1.202 | 2.345 | **0.021** |
| iHPT | 44 | 0.261 | 1.575 | 0.927 | 0.357 | 43 | 0.584 | 1.830 | 1.835 | 0.070 |
| iPPT | 43 | -0.365 | 1.420 | -1.377 | 0.172 | 44 | -0.467 | 1.331 | -1.862 | 0.066 |
| iMPT | 45 | 1.349 | 1.629 | 4.734 | **0.000** | 45 | 1.706 | 1.440 | 6.529 | **0.000** |
| iMPS | 45 | 1.362 | 1.620 | 4.797 | **0.000** | 45 | 1.527 | 1.433 | 5.863 | **0.000** |
| iWUR | 43 | 0.030 | 1.154 | 0.128 | 0.898 | 44 | -0.231 | 0.885 | -1.146 | 0.255 |
| iMDT | 45 | -0.490 | 1.366 | -1.943 | 0.055 | 45 | -0.535 | 1.504 | -1.989 | 0.050 |
| iVDT | 45 | 0.357 | 1.280 | 1.474 | 0.144 | 44 | 0.397 | 1.063 | 1.802 | 0.075 |
| sdCDT | 44 | -0.464 | 1.692 | -1.565 | 0.121 | 43 | -0.154 | 1.372 | -0.597 | 0.552 |
| sdWDT | 44 | -0.970 | 1.622 | -3.376 | 0.001 | 43 | -0.519 | 1.557 | -1.841 | 0.069 |
| sdTSL | 44 | -0.879 | 1.569 | -3.134 | 0.002 | 43 | -0.573 | 1.640 | -1.957 | 0.054 |
| sdCPT | 44 | 1.022 | 1.759 | 3.349 | 0.001 | 43 | 0.584 | 1.394 | 2.232 | 0.028 |
| sdHPT | 44 | -0.199 | 1.518 | -0.727 | 0.469 | 43 | -0.057 | 1.938 | -0.170 | 0.865 |
| sdPPT | 42 | 0.042 | 0.926 | 0.202 | 0.841 | 44 | 0.002 | 1.241 | 0.007 | 0.995 |
| sdMPT | 45 | 0.117 | 2.138 | 0.332 | 0.740 | 45 | -0.019 | 1.368 | -0.077 | 0.939 |
| sdMPS | 45 | -0.506 | 1.837 | -1.623 | 0.108 | 45 | 0.005 | 1.487 | 0.019 | 0.985 |
| sdWUR | 42 | 0.095 | 1.247 | 0.386 | 0.701 | 44 | 0.230 | 0.976 | 1.090 | 0.279 |
| sdMDT | 45 | -1.198 | 2.610 | -2.874 | 0.005 | 45 | 0.108 | 2.301 | 0.287 | 0.774 |
| sdVDT | 45 | -0.312 | 1.007 | -1.476 | 0.144 | 44 | -0.279 | 1.082 | -1.254 | 0.213 |

CPSP: central post-stroke pain; NPSS: non–pain sensory stroke; QST; Quantitative Sensory Testing; CDT, cold detection threshold; CPT, cold pain threshold; HPT, heat pain threshold; MDT, mechanical detection threshold; MPS, mechanical pain sensitivity; MPT,mechanical pain threshold; PPT, pressure pain threshold; TSL, temperature sensory limen; VDT, vibration detection threshold; WDT, warm detection threshold; WUR, wind- up ratio; QST parameters on the contralesional side are indicated with “_c_” (e.g., _c_CDT), on the ipsilesional with “_i_” (e.g., _i_CDT). Side-to-side differences are indicated with “_sd_” (e.g., _sd_CDT).

Statistical comparisons of QST values to DFNS reference values. A total of 45 NPSS patients were included the N column indicates the number of participants for each QST variable. P-values were obtained by comparing the NPSS group’s Z-standardised QST mean with a virtual reference population mean as suggested by Magerl et al 2010 assuming a mean of 0 and SD = 1. Alpha level was 0.05, p-values are not Bonferroni-corrected. Blue boxes indicate a loss of function and orange boxes a gain of function.

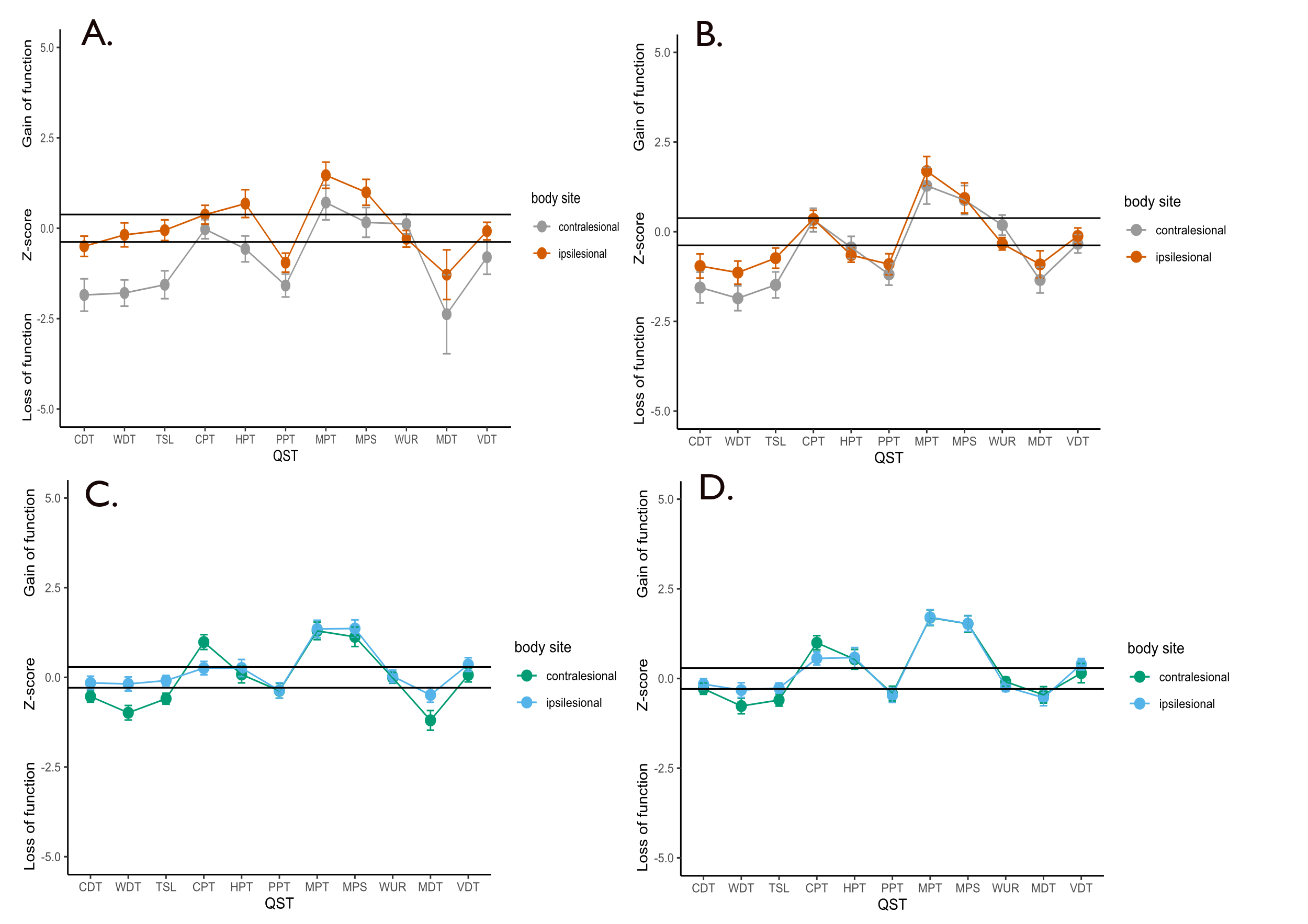
**Figure S1: Abnormal QST parameters in the acute and chronic phase for CPSP and NPSS patients**

CPSP: central post-stroke pain; NPSS: non–pain sensory stroke; QST; Quantitative Sensory Testing; CDT, cold detection threshold; CPT, cold pain threshold; HPT, heat pain threshold; MDT, mechanical detection threshold; MPS, mechanical pain sensitivity; MPT, mechanical pain threshold; PPT, pressure pain threshold; TSL, temperature sensory limen; VDT, vibration detection threshold; WDT, warm detection threshold; WUR, wind- up ratio

A. CPSP acute. B. CPSP chronic. C. NPSS acute. D. NPSS chronic. Mean and the standard error of the mean (SEM) are plotted above. QST parameters that are significantly different from the reference collective for each group are outside the confidence intervals. The confidence interval for the CPSP and NPSS groups was adjusted for the numbers of observations by dividing the standard confidence interval through the root of the number of observations ((±1.96/ √ (number of observations) for two-sided confidence intervals and 1.64/√ (number of observations) for one-sided confidence intervals (VDT). The number of observations was n = 26 for CPSP and n = 49 for NPSS, indicating a deviation from the reference values (for the respective group) if the mean QST values lie outside the margin of ±0.38/ ±0.32 for CPSP patients and outside the marge of ±0.29/ ±0.23 for NPSS patients.

**Table S6: Frequency of different QST combinations of abnormal values in CPSP and NPSS patients in the acute phase before pain onset.**

CPSP: central post-stroke pain; NPSS: non–pain sensory stroke; QST; Quantitative Sensory Testing;

| **Loss** | **Gain** | | | | |
| --- | --- | --- | --- | --- | --- |
|  | **No gain** | **Thermal only** | **Mechanical only** | **Mixed** | All |
| **CPSP** |  |  |  |  |  |
| **No loss** | 0% | 2 (11%) | 2 (11%) | 0% | 4 (21%) |
| **Thermal only** | 3 (16%) | 0% | 1 (5%) | 1 (5%) | 5 (26%) |
| **Mechanical only** | 1(5%) | 1 (5%) | 0% | 0% | 2 (11%) |
| **Mixed** | 2 (11%) | 1(5%) | 4 (21%) | 1 (5%) | 8 (42%) |
| All | 6 (32%) | 4 (21%) | 7 (37%) | 2 (11%) | 19 (100%) |
| **NPSS** |  |  |  |  |  |
| **No loss** | 3 (7%) | 2 (5%) | 5 (12%) | 4 (9%) | 14 (33%) |
| **Thermal only** | 0% | 0% | 4 (9%) | 2 (5%) | 6 (14%) |
| **Mechanical only** | 2 (5%) | 2 (5%) | 3 (7%) | 4 (9%) | 11 (26%) |
| **Mixed** | 3 (7%) | 0% | 3 (7%) | 6 (14%) | 12 (28%) |
| All | 8 (19%) | 4 (9%) | 15 (35%) | 16 (37%) | 43 (100%) |

Loss: No loss = no loss of function in detection; thermal only = thermal loss of function in the parameters contralesional CDT, WDT or the side difference; mechanical only = loss of function in contralesional MDT, VDT, or side difference. Mixed = loss of function in thermal (contralesional CDT or WDT or side difference) and mechanical parameters (contralesional MDT or VDT or side difference).

Gain: No gain = no hyperalgesia; thermal only, hyperalgesia in contralesional CPT or HPT or the side difference; mechanical only, hyperalgesia in contralesional MPT, MPS, allodynia or the side difference; mixed, gain of function in both mechanical and thermal parameters. The number of patients (%), percentage of patients with condition rounded up to the nearest whole number. CPSP, central post stroke pain; NPSS, non-pain sensory stroke. Loss of function was defined z scores < -1.96 and gain of function as z scores > 1.96.

**Table S7. Frequency of different QST combinations of abnormal values in CPSP and NPSS patients in the chronic phase after pain onset.**

| **Loss** | **Gain** | | | | |
| --- | --- | --- | --- | --- | --- |
|  | **No gain** | **Thermal only** | **Mechanical only** | **Mixed** | All |
| **CPSP** |  |  |  |  |  |
| **No loss** | 2(11%) | 2(11%) | 1(5%) | 0% | 5(26%) |
| **Thermal only** | 0% | 0% | 3(16%) | 1(5%) | 4(21%) |
| **Mechanical only** | 2(11%) | 0% | 1(5%) | 1(5%) | 4(21%) |
| **Mixed** | 2(11%) | 0% | 3(16%) | 1(5%) | 6(32%) |
| All | 6(32%) | 2(11%) | 8(42%) | 3(16%) | 19(100%) |
| **NPSS** |  |  |  |  |  |
| **No loss** | 4(10%) | 2(5%) | 6(15%) | 6(15%) | 18(44%) |
| **Thermal only** | 3(7%) | 0% | 0% | 1(2%) | 4(10%) |
| **Mechanical only** | 2(5%) | 4(10%) | 4(10%) | 3(7%) | 13(32%) |
| **Mixed** | 1(2%) | 0% | 3(7%) | 2(5%) | 6(15%) |
| All | 10(24%) | 6(15%) | 13(32%) | 12(29%) | 41(100%) |

CPSP, central post-stroke pain; NPSS, non–pain sensory stroke; QST, Quantitative Sensory Testing

Loss: No loss, no loss of function in detection; thermal only, thermal loss of function in the parameters contralesional CDT, WDT or the side difference; mechanical only, loss of function in contralesional MDT, VDT, or side difference. Mixed, loss of function in thermal (contralesional CDT or WDT or side difference) and mechanical parameters (contralesional MDT or VDT or side difference).

Gain: No gain, no hyperalgesia; thermal only gain, hyperalgesia in contralesional CPT or HPT or the side difference; mechanical only, hyperalgesia in contralesional MPT, MPS, allodynia or the side difference; mixed, gain of function in both mechanical and thermal parameters. The number of patients (%), percentage of patients with condition rounded up to the nearest whole number. CPSP, central post stroke pain; NPSS, non-pain sensory stroke. Loss of function was defined z scores < -1.96 and gain of function as z scores > 1.96.

**Table S8: Longitudinal analysis of QST changes over time in the acute (before pain) and chronic (with pain) setting**

| **A.**  **Contrast**  *p* F [df1, df2] |  |  |  |  |  |  |  |  |  |  |  |
| --- | --- | --- | --- | --- | --- | --- | --- | --- | --- | --- | --- |
|  | **cCDT** | **cWDT** | **cTSL** | **cCPT** | **cHPT** | **cPPT** | **cMPT** | **cMPS** | **cWUR** | **cMDT** | **cVDT** |
| Group | **0.016**  6.5[1,31] | **0.015**  6.5[1,40] | **0.027**  5.3[1,34] | **0.003**  9.4[1,50] | **0.005**  8.8[1,52] | **0.002**  10.4[1,51] | 0.443  0.6[1,33] | 0.081  3.2[1,51] | 0.343  0.9[1,49] | 0.138  2.3[1,49] | **0.010**  7.4[1,39] |
| Time | 0.225  1.5[1,32] | 0.779  0.1[1,43] | 0.702  0.1[1,36] | 0.524  0.4[1,45] | 0.171  1.9[1,51] | 0.660  0.2[1,35] | 0.285  1.2[1,37] | 0.274  1.2[1,32] | 0.522  0.4[1,38] | 0.146  2.2[1,38] | 0.342  0.9[1,42] |
| Group*Time | 0.725  0.1[1,32] | 0.261  1.3[1,43] | 0.868  0.03[1,36] | 0.411  0.7[1,45] | 0.779  0.1[1,51] | 0.491  0.5[1,35] | 0.786  0.1[1,37] | 0.413  0.7[1,32] | 0.574  0.3[1,38] | 0.671  0.2[1,38] | 0.895  0.0[1,42] |
| **B.** | **iCDT** | **iWDT** | **iTSL** | **iCPT** | **iHPT** | **iPPT** | **iMPT** | **iMPS** | **iWUR** | **iMDT** | **iVDT** |
| Group | 0.080  3.2[1,43] | 0.177  1.9[1,48] | 0.802  0.1[1,43] | 0.773  0.1[1,46] | 0.707  0.1[1,52] | 0.250  1.4[1,52] | 0.808  0.06[1,40] | 0.439  0.6[1,52] | 0.487  0.5[1,46] | 0.354  0.9[1,50] | **0.006**  8.5[1,37] |
| Time | 0.287  1.6[1,45] | **0.004**  9.0[1,52] | **0.021**  5.7[1,42] | 0.305  1.1[1,47] | 0.126  2.4[1,48] | 0.994  0.0[1,35] | 0.284  1.2[1,47] | 0.832  0.05[1,39] | 0.507  0.4[1,44] | 0.732  0.1[1,42] | 0.862  0.03[1,34] |
| Group*Time | 0.515  0.4[1,45] | **0.029**  5.0[1,52] | 0.090  3.0[1,42] | 0.387  0.8[1,47] | **0.014**  6.5[1,48] | 0.840  0.0[1,35] | 0.866  0.03[1,47] | 0.925  0.01[1,39] | 0.411  0.7[1,44] | 0.932  0.01[1,42] | 0.743  0.1[1,34] |
| **C.** | **sdCDT** | **sdWDT** | **sdTSL** | **sdCPT** | **sdHPT** | **sdPPT** | **sdMPT** | **sdMPS** | **sdWUR** | **sdMDT** | **sdVDT** |
| Group | 0.157  2.1[1,37] | 0.145  2.2[1,38] | 0.092  3.0[1,35] | **0.0003**  14.8[1,51] | **0.011**  7.3[1,33] | 0.114  2.6[1,33] | 0.171  1.9[1,41] | 0.268  1.3[1,30] | 0.061  3.7[1,47] | 0.076  3.3[1,43] | 0.123  2.5[1,32] |
| Time | **0.030**  5.0[1,44] | 0.074  3.3[1,48] | 0.121  2.5[1,36] | 0.693  0.2[1,51] | **0.011**  7.1[1,40] | 0.758  0.1[1,34] | 0.857  0.03[1,39] | 0.062  3.8[1,30] | 0.443  0.6[1,44] | 0.483  0.5[1,34] | 0.373  0.8[1,35] |
| Group*Time | 0.360  0.9[1,44] | 0.434  0.6[1,48] | 0.176  1.9[1,36] | 0.064  3.6[1,51] | **0.046**  4.2[1,40] | 0.761  0.1[1,34] | 0.646  0.2[1,39] | 0.486  0.5[1,30] | 0.284  1.2[1,44] | 0.251  1.4[1,34] | 0.654  0.2[1,35] |

QST, Quantitative Sensory Testing; CDT, cold detection threshold; CPT, cold pain threshold; HPT, heat pain threshold; MDT, mechanical detection threshold; MPS, mechanical pain sensitivity; MPT, mechanical pain threshold; PPT, pressure pain threshold; TSL, temperature sensory limen; VDT, vibration detection threshold; WDT, warm detection threshold; WUR, wind- up ratio; QST on the contralesional side is indicated with “c” (e.g., cCDT), on the ipsilesional with “i” (e.g., iCDT). Side-to-side differences are indicated with “sd” (e.g., sdCDT).

Robust ANOVA with 20% trimmed means was conducted to investigate whether there is a group by time interaction between CPSP and NPSS patients across the acute and chronic settings. P values are reported above as well as F values and degrees of freedom (rounded up to the nearest whole number). Post-hoc tests were conducted with 10000 permutations for all significant interactions.

**Table S9: Multiple logistic regression models**

| **Diagnosis** |  | **NPSS** | **CPSP** | **OR (univariable)** | **OR (multivariable full)** | **OR (multivariable)** |
| --- | --- | --- | --- | --- | --- | --- |
| NIHSS | Mean (SD) | 1.9 (2.0) | 4.2 (4.3) | 1.38 (1.09-1.89, p=0.023) | 1.69 (1.01-3.42, p=0.074) | 1.17 (0.84-1.66, p=0.326) |
| Sex | Male | 33 (78.6) | 9 (21.4) | - | - | - |
|  | Female | 12 (52.2) | 11 (47.8) | 3.36 (1.13-10.41, p=0.031) | 14.40 (0.76-726.97, p=0.109) | 5.03 (0.80-42.97, p=0.100) |
| sdCDT | Mean (SD) | -0.5 (1.7) | -1.7 (2.5) | 0.73 (0.54-0.96, p=0.031) | 2.01 (0.55-10.98, p=0.325) | - |
| sdTSL | Mean (SD) | -0.9 (1.6) | -2.3 (2.4) | 0.68 (0.49-0.91, p=0.013) | 0.42 (0.09-1.37, p=0.185) | - |
| sdCPT | Mean (SD) | 1.0 (1.8) | -0.7 (1.7) | 0.57 (0.37-0.80, p=0.003) | 0.51 (0.18-1.04, p=0.104) | 0.54 (0.28-0.89, p=0.029) |
| sdHPT | Mean (SD) | -0.2 (1.5) | -1.5 (2.4) | 0.68 (0.48-0.92, p=0.017) | 0.82 (0.32-1.96, p=0.663) | 0.59 (0.30-1.03, p=0.082) |
| sdMPT | Mean (SD) | 0.1 (2.1) | -1.2 (2.5) | 0.78 (0.60-0.98, p=0.044) | 0.86 (0.45-1.53, p=0.614) | - |
| sdMPS | Mean (SD) | -0.5 (1.8) | -1.9 (3.4) | 0.78 (0.60-0.98, p=0.047) | 0.93 (0.40-2.06, p=0.840) | - |
| sdWUR | Mean (SD) | 0.1 (1.2) | 0.8 (1.1) | 1.74 (1.05-3.11, p=0.042) | 2.01 (0.95-6.54, p=0.119) | - |
| cCDT | Mean (SD) | -0.5 (1.1) | -1.8 (2.0) | 0.55 (0.35-0.80, p=0.004) | 0.46 (0.09-1.74, p=0.275) | 0.57 (0.27-1.05, p=0.088) |
| cWDT | Mean (SD) | -1.0 (1.4) | -1.8 (1.6) | 0.68 (0.46-0.98, p=0.047) | 2.47 (0.34-30.03, p=0.392) | - |
| cTSL | Mean (SD) | -0.6 (1.0) | -1.6 (1.7) | 0.56 (0.34-0.85, p=0.012) | 0.43 (0.01-6.95, p=0.558) | - |
| cCPT | Mean (SD) | 1.0 (1.4) | -0.0 (1.2) | 0.55 (0.33-0.84, p=0.010) | 0.48 (0.10-1.65, p=0.279) | - |
| cPPT | Mean (SD) | -0.4 (1.3) | -1.6 (1.4) | 0.51 (0.30-0.78, p=0.005) | 0.23 (0.04-0.70, p=0.034) | 0.24 (0.07-0.56, p=0.006) |

Full multivariable model, number in dataframe = 65, Number in model = 56, Missing = 9, AIC = 60.5, C-statistic = 0.944, H&L = Chi-sq (8) 5.57 (*P* = 0.696).

Reduced multivariable model, number in dataframe = 65, Number in model = 60, Missing = 5, AIC = 51.5, C-statistic = 0.927, H&L = Chi-sq (8) 6.20 (*P* = 0.625)

CPSP: central post-stroke pain; NPSS: non–pain sensory stroke; QST; Quantitative Sensory Testing; National Institutes of Health Stroke Scale: NIHSS; CDT, cold detection threshold; CPT, cold pain threshold; HPT, heat pain threshold; MDT, mechanical detection threshold; MPS, mechanical pain sensitivity; MPT,mechanical pain threshold; PPT, pressure pain threshold; TSL, temperature sensory limen; VDT, vibration detection threshold; WDT, warm detection threshold; WUR, wind- up ratio; QST on the contralesional side is indicated with “c” (e.g., cCDT), on the ipsilesional with “i” (e.g., iCDT). Side-to-side differences are indicated with “sd” (e.g., sdCDT).

**Figure S2: QST parameters in the acute phase after stroke for CPSP and NPSS patients**

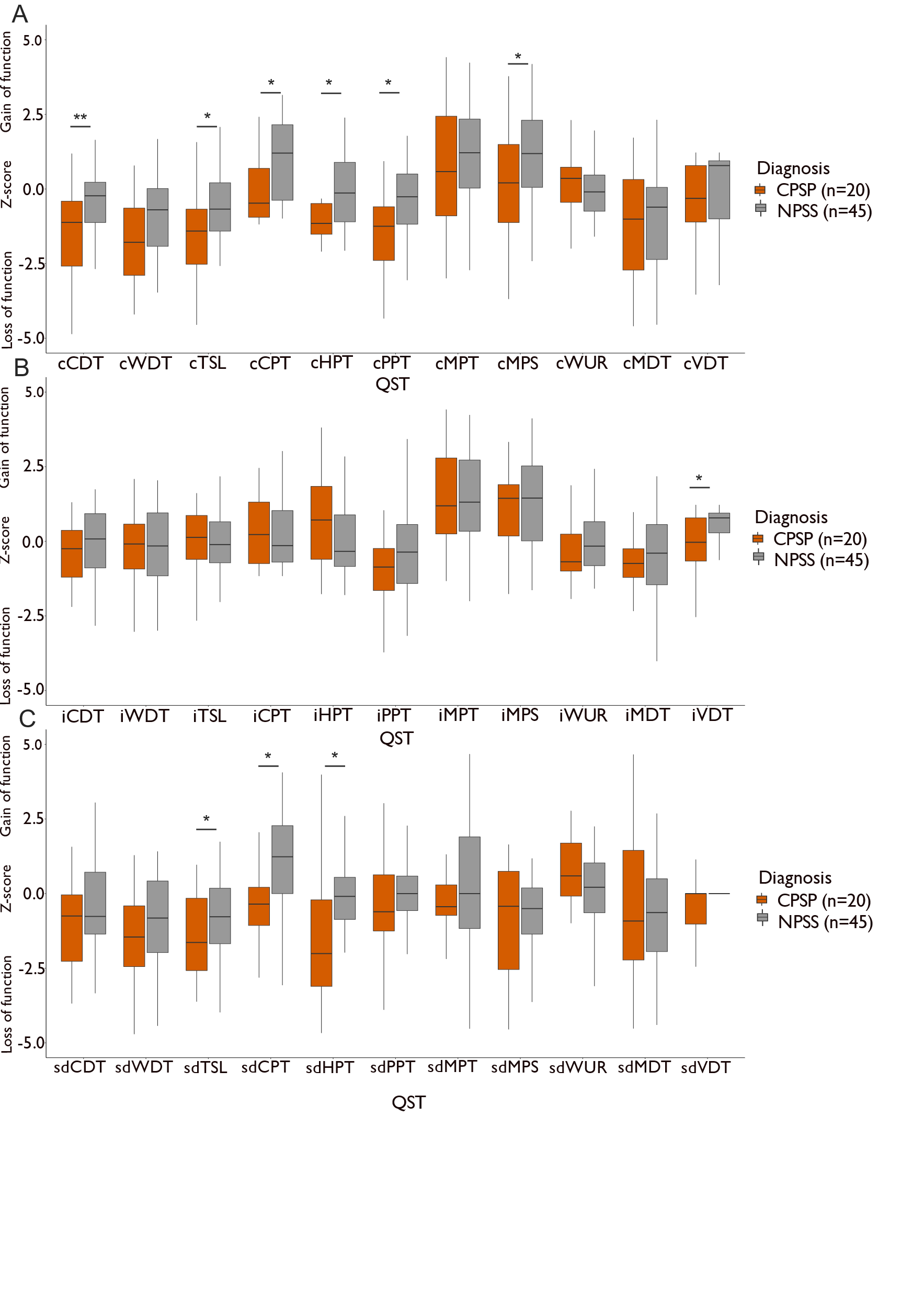

CPSP: central post-stroke pain; NPSS: non–pain sensory stroke; QST; Quantitative Sensory Testing; National Institutes of Health Stroke Scale: NIHSS; CDT, cold detection threshold; CPT, cold pain threshold; HPT, heat pain threshold; MDT, mechanical detection threshold; MPS, mechanical pain sensitivity; MPT,mechanical pain threshold; PPT, pressure pain threshold; TSL, temperature sensory limen; VDT, vibration detection threshold; WDT, warm detection threshold; WUR, wind- up ratio; QST on the contralesional side is indicated with “c” (e.g., cCDT), on the ipsilesional with “i” (e.g., iCDT). Side-to-side differences are indicated with “sd” (e.g., sdCDT).

**A.** Box plot of QST parameters on the contralesional side in the acute phase after stroke. **B.** QST parameters on the ipsilesional side. **C.** Side to side differences of QST parameters. Mann-Whitney U tests performed, and significant results are marked with an * *P* < 0.05 uncorrected, ** *P* < 0.05, Bonferroni corrected. At the individual level a gain of function is determined when the z score is 2SD above 0 and a loss of function when the z score is 2SD below 0.

**Figure S3: QST parameters in the chronic phase after stroke for CPSP and NPSS patients**

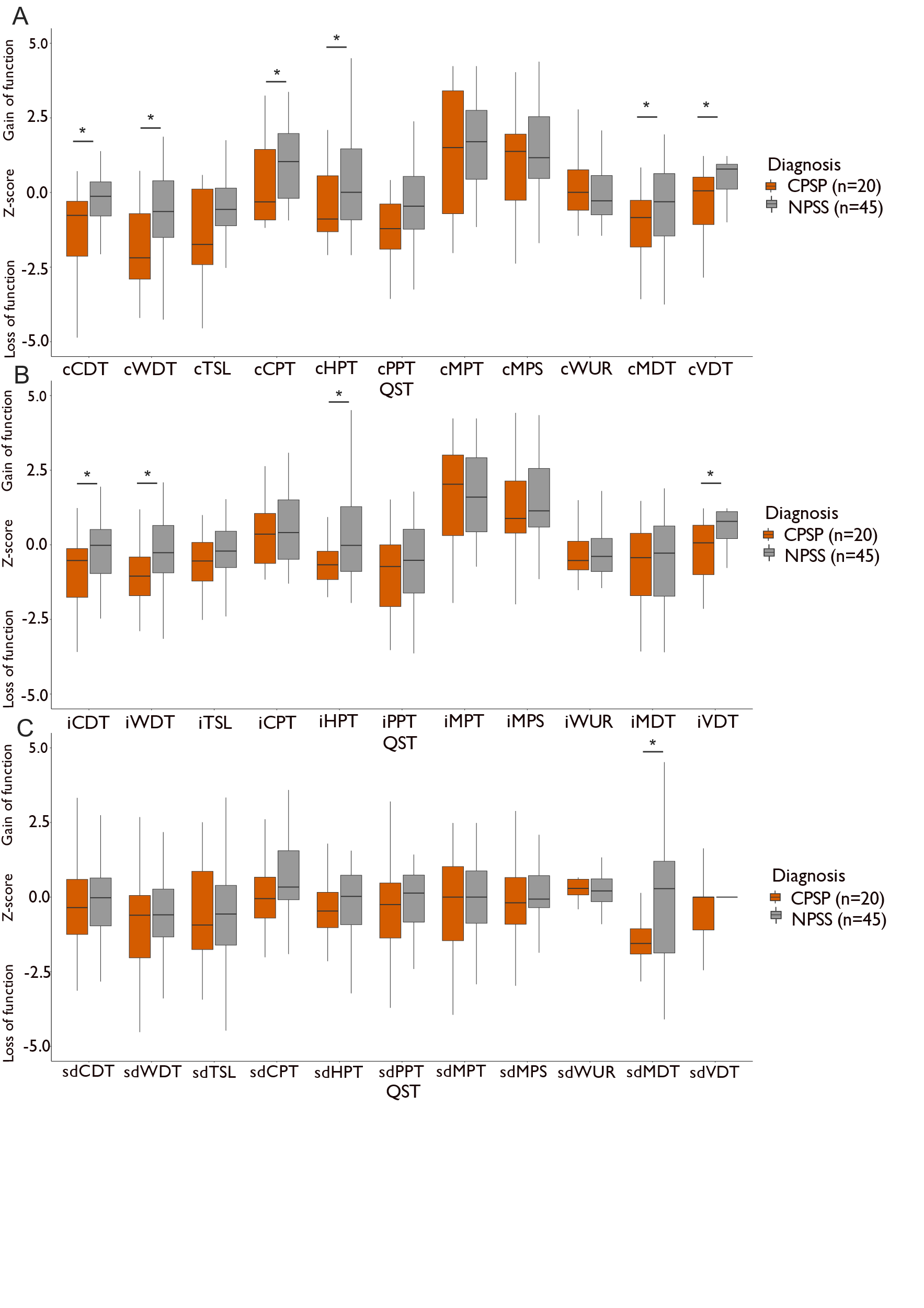

CPSP: central post-stroke pain; NPSS: non–pain sensory stroke; QST; Quantitative Sensory Testing; National Institutes of Health Stroke Scale: NIHSS; CDT, cold detection threshold; CPT, cold pain threshold; HPT, heat pain threshold; MDT, mechanical detection threshold; MPS, mechanical pain sensitivity; MPT,mechanical pain threshold; PPT, pressure pain threshold; TSL, temperature sensory limen; VDT, vibration detection threshold; WDT, warm detection threshold; WUR, wind- up ratio; QST on the contralesional side is indicated with “c” (e.g., cCDT), on the ipsilesional with “i” (e.g., iCDT). Side-to-side differences are indicated with “sd” (e.g., sdCDT)

**A.** Box plot of QST on the contralesional side in the chronic phase after stroke. **B.** QST on the ipsilesional side. **C.** QST side to side differences. Mann-Whitney U tests performed and significant results are marked with an * *P* < 0.05 uncorrected, ** *P* < 0.05, Bonferroni corrected. At the individual level a gain of function is determined when the z score is 2SD above 0 and a loss of function when the z score is 2SD below 0.
